# Genetic variants risk assessment for Long QT Syndrome through machine learning and multielectrode array recordings

**DOI:** 10.1101/2025.03.25.25324187

**Authors:** Aleksandr Khudiakov, Manuela Mura, Federica Giannetti, Vladislav Leonov, Chiara Alberio, Marem Eskandr, Paul A Brink, Lia Crotti, Massimiliano Gnecchi, Peter J Schwartz, Luca Sala

**Affiliations:** Istituto Auxologico Italiano IRCCS, Center for Cardiac Arrhythmias of Genetic Origin and Laboratory of Cardiovascular Genetics, Milan, Italy; Translational Cardiology Unit, Fondazione IRCCS Policlinico San Matteo, Pavia, Italy; Department of Surgery, Dentistry, Pediatrics and Gynecology, Cardiovascular Science, The University of Verona, Policlinico G.B. Rossi, Verona, Italy; Department of Biotechnology and Biosciences, University of Milano – Bicocca, Italy; Department of Medicine, University of Stellenbosch, Tygerberg, South Africa; Department of Medicine and Surgery, University of Milano-Bicocca, Milan, Italy; Department of Molecular Medicine, Unit of Cardiology, University of Pavia, Pavia, Italy

**Keywords:** Precision medicine, genetic variants, risk assessment, machine learning, long QT syndrome, hiPSC-derived cardiomyocytes, multielectrode arrays, proarrhythmic drugs, safety pharmacology

## Abstract

**Background:** Long QT syndrome (LQTS) is a life-threatening genetic disorder characterized by prolonged QT intervals on electrocardiograms. Congenital forms are mostly associated with variants in the *KCNQ1* and *KCNH2* genes. Among pathogenic or likely pathogenic (P/LP) variants, some are associated with a significantly higher incidence of cardiac events compared to others. While therapies have significantly reduced mortality, some patients are unresponsive or intolerant to therapy, perpetuating their arrhythmic risk, including sudden cardiac death. Current approaches for risk stratification are insufficient, highlighting the critical need for more accurate identification and management of patients carrying high risk genetic variants.

**Objectives:** To develop a refined risk stratification model for P/LP variants by applying machine learning classification to electrophysiological data measured in human induced pluripotent stem cell-derived cardiomyocytes (hiPSC-CMs).

**Methods:** Eleven patient-specific hiPSC lines carrying six P/LP variants in *KCNQ1* or *KCNH2* were differentiated to cardiomyocytes (hiPSC-CMs). Electrophysiological responses from multielectrode array recordings at baseline and after application of selective ion channel blockers or pro-arrhythmic compounds were used to train a machine learning model to classify variant-specific risk levels based on *in vitro* electrophysiological readouts.

**Results:** Our findings revealed a correlation between variant risk level, hiPSC-CM electrophysiological profiles, and drug responses. The machine learning classifier, trained on multielectrode array recordings, achieved 89% accuracy in classification of P/LP genetic variants according to the associated risk levels.

**Conclusions:** This study demonstrates that integrating hiPSC-CM electrophysiological profiling with machine learning provides a robust method to improve variant-specific risk stratification for LQTS patients.

**Clinical Perspectives:** *Clinical Aspects:* Understanding which patients may be at risk of cardiac events or sudden cardiac death is crucial to implement appropriate preventive measures. This study leverages patient-specific *in vitro* models and machine learning to improve the risk stratification of pathogenic/likely pathogenic variants associated with LQTS, better supporting clinical decisions related to risk assessment and management of LQTS patients. This scalable approach can be implemented across multiple centres, enhancing the risk stratification of LQTS variants beyond what is currently possible when clinical data are limited.

*Translational Outlook:* Machine learning-based variant risk stratification is a novel approach for integrating hiPSC-CM-derived electrophysiological data into clinical workflows. While this study demonstrates the feasibility of our approach, further research is required to validate these findings across larger and more diverse patient cohorts. Additionally, efforts to standardize the pipeline and adapt it for multicentric implementation are necessary. Graphical Abstract

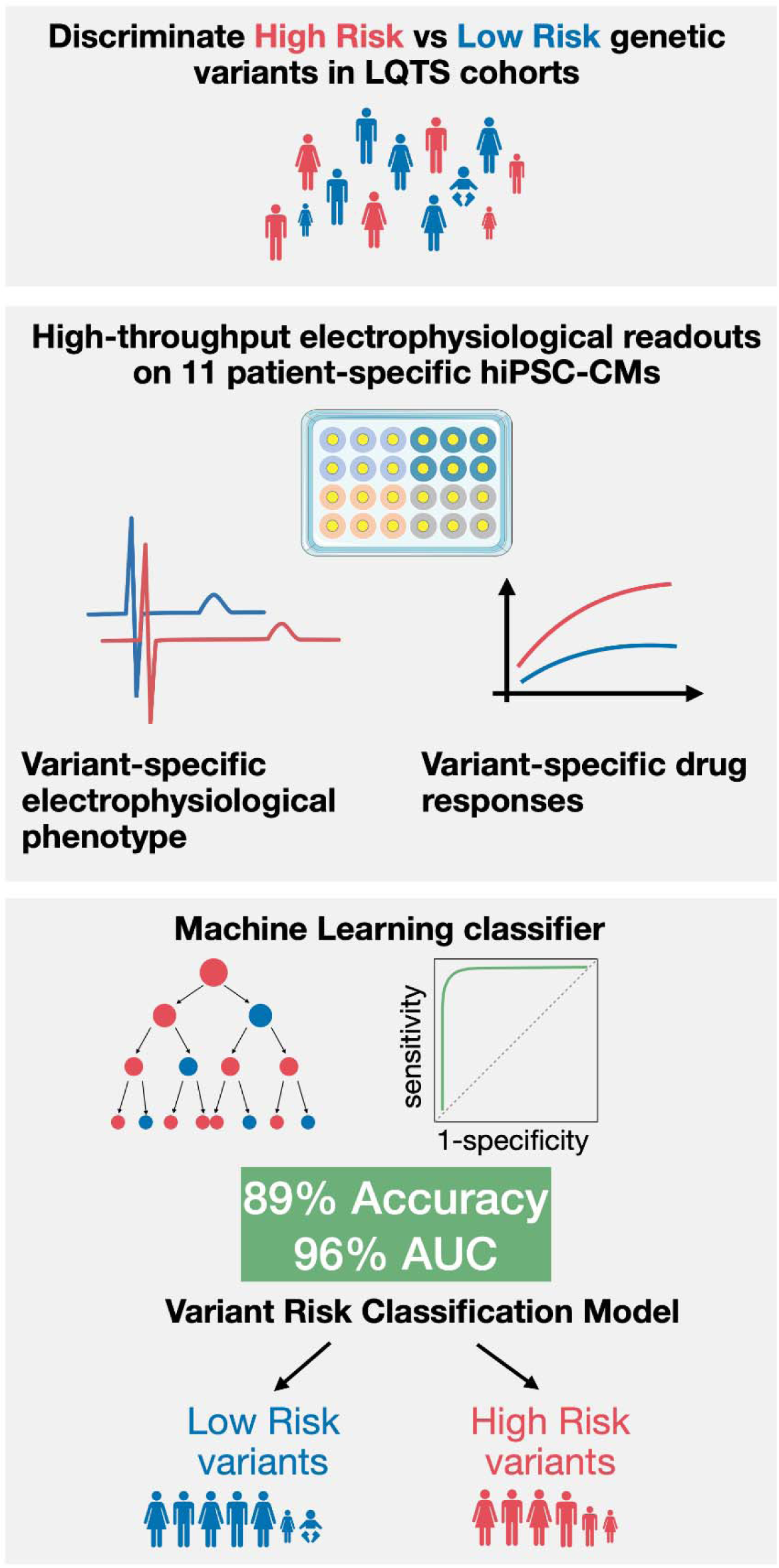

**Highlights:**

- Discriminating LQTS patients at high or low risk for sudden death is a clinical challenge.
- Improved stratification of pathogenic/likely pathogenic variants is achievable through machine learning classification on in vitro electrophysiological data.
- Integration of the clinical workflow with data from patient-specific in vitro models will enhance risk stratification.

## Introduction

The long QT syndrome (LQTS) is a life-threatening disease of genetic origin characterized by a prolonged QT interval on the electrocardiogram, by propensity to lethal arrhythmias especially under stress, and by an increased sensitivity to drugs affecting cardiac repolarization through block of the I_Kr_ potassium current ^1–4^. Appropriate therapies based on beta-blockers ^5^, left cardiac sympathetic denervation ^6,7^ and sodium channel blockers ^8,9^ have effectively reduced the risk for patients to develop potentially-lethal ventricular arrhythmias or sudden cardiac death (SCD), limiting the use of an implantable cardioverter defibrillator only to selected patients ^5,10^; one of the main challenges for clinicians dealing with LQTS is the identification, clinical management and protection of patients who are more at risk of developing SCD.

Advancements in next-generation sequencing have led to the discovery of numerous variants in genes encoding cardiac ion channels, subunits, or associated proteins implicated in LQTS ^11,12^. While these insights have guided gene-specific patient management ^8^, current genetic interpretation guidelines ^13^ often do not help identify patients at higher risk, with a large number of variants classified as variants of uncertain significance (VUS). Even variants classified as pathogenic or likely pathogenic (P/LP) according to the American College of Medical Genetics and Genomics (ACMG) guidelines may exhibit divergent risks of life-threatening cardiac events, including SCD ^14,15^, requiring profoundly different clinical management.

Accurate clinical risk assessment for a variant requires data from many affected individuals and this is not feasible for most of the variants, limiting precision medicine and hampering the development of variant-specific therapies ^16,17^. The classification of variant pathogenicity through *in vitro* or *in vivo* studies offers a promising solution to address this limitation ^18,19^. Functional studies using patient-specific cardiomyocytes derived from human induced pluripotent stem cells (hiPSC-CMs) further refined this, providing high-quality functional data on the effect of the variants on action potentials, calcium transients or contractility. hiPSC-CM-based models have been particularly useful for disease modeling of congenital or acquired cardiac disorders such as LQTS ^20–24^, Jervell and Lange-Nielsen syndrome (JLNS) ^25^, Timothy syndrome ^26^, cardiomyopathies ^27–29^, congenital heart defects ^30^, drug testing ^31–33^ and drug repurposing ^34^. In the present study, using hiPSC-CMs from eleven patients, we focused on six well-characterized P/LP variants from the two most prevalent LQTS subtypes, LQT1 and LQT2, which together account for 90% of cases. These variants were identified in individuals with clinically heterogeneous presentations, from normal to prolonged QT intervals and displaying low or high incidence of life-threatening cardiac events per variant (defined here as Low and High Risk P/LP variants).

Using patient-derived hiPSC-CMs and high-throughput multielectrode arrays (MEA), we investigated i) whether hiPSC-CMs carrying P/LP variants with differing clinical severities exhibit distinct drug responses *in vitro*; ii) whether *in vitro* phenotypes match clinical records and could be used for a more accurate variant-associated risk stratification; iii) whether accurate P/LP variant risk stratification can be automated through machine learning on *in vitro* readouts.

## Methods

### Ethical statement

This study was conducted in accordance with the Declaration of Helsinki and the ethics committee of Istituto Auxologico Italiano IRCCS gave ethical approval for this work (Approval number: 2020_10_20_07). Appropriate informed consents were obtained from all donors.

### Patient-specific hiPSC lines included in the study

A total of eleven hiPSC lines were utilized in this study. HiPSC lines were derived from LQT1, JLNS, and LQT2 patients carrying the variants in *KCNQ1* or *KCNH2* gene (See Table 1 for detailed information). The line carrying the *KCNH2* p.A561V variant was provided by Joseph C. Wu, MD, PhD, at the Stanford Cardiovascular Institute. As a wild-type control (WT), the WTC-11 line, generated from a healthy subject, was obtained from the Coriell Institute for Medical Research (catalog No. GM25256) and used as a bona fide wild-type reference ^35^. Detailed characterization of each hiPSC line, including genetic and functional properties, is provided in Supplemental Table 1 and Supplemental Figure 1 and Supplemental Figure 2.

**Table 1.**
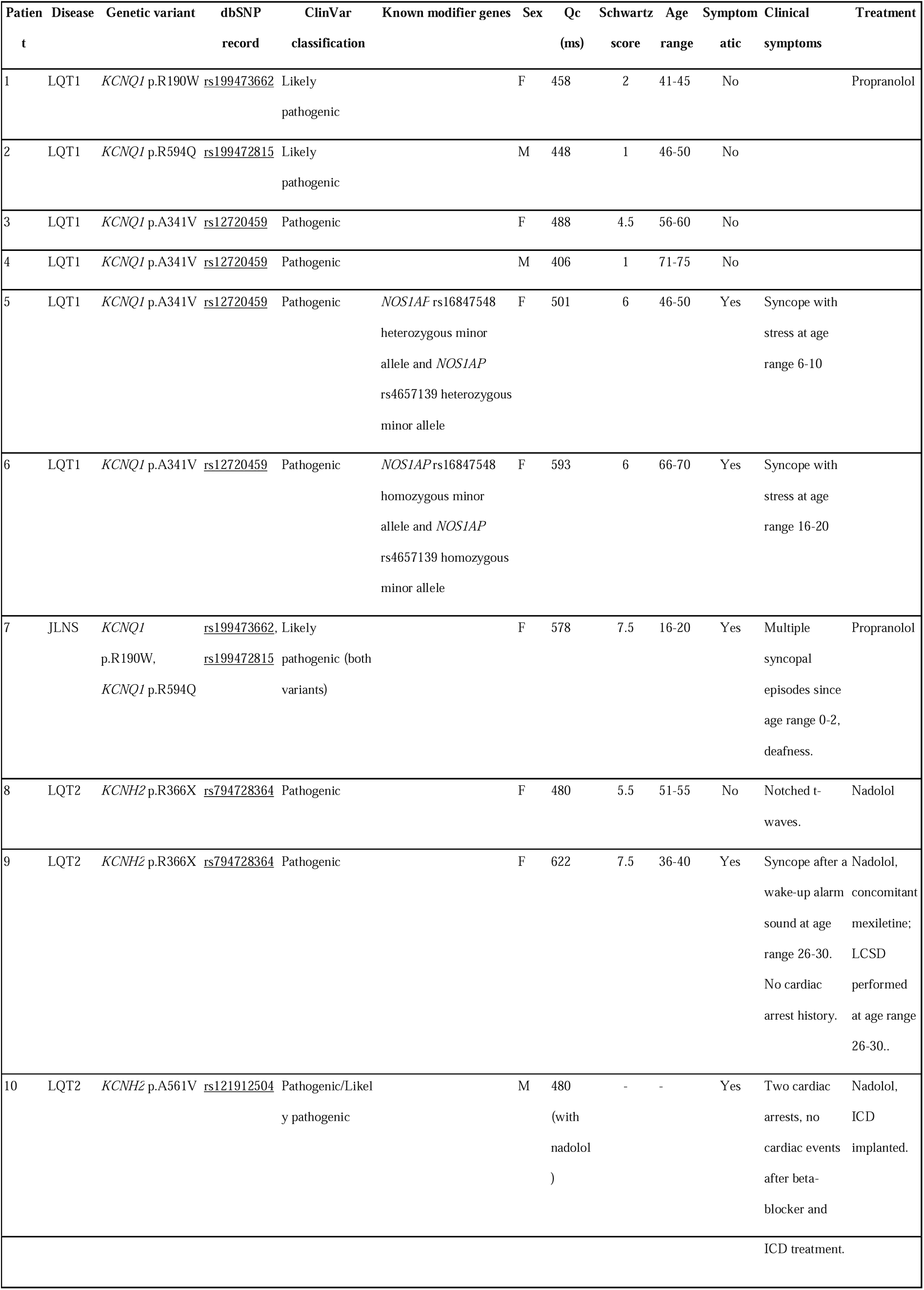
Clinical characteristics of the patient cohort included in the study.

### Cardiac events frequency assessment

Clinical data on documented cardiac events (syncope, sustained ventricular tachycardia, appropriate ICD shock, sudden cardiac arrest, sudden cardiac death), QTc values and Schwartz scores (when reported) were extracted from our internal database of LQTS patients, The Human Gene Mutation Database (HGMD)^36^ and Variant Browser database ^37,38^. The frequency of cardiac events was calculated as the proportion of patients with a history of such events relative to the total number of patients carrying the specific variant.

### hiPSC culture and differentiation to hiPSC-CMs

HiPSCs were cultured on multiwell plates coated with recombinant human vitronectin (Gibco) in Essential 8 Flex medium kit (Gibco). HiPSCs were plated on Matrigel (Corning) coated plates for cardiac differentiation. hiPSCs were differentiated to hiPSC-CMs following a small-molecule based protocol ^39^, purified through glucose starvation ^40^, starting from day 7, and cryopreserved in Bambanker (Nippon Genetics) at days 9-16. For the subsequent experiments hiPSC-CMs were thawed, replated at low density and expanded as previously reported ^41^. A list of main reagents used in the study is reported in Supplemental Table 2. Data were collected from at least three independent differentiations of each hiPSC line for each experiment.

### Patch-clamp

For patch clamp experiments, metabolic maturation of hiPSC-CMs was performed as previously described ^42^. Action potentials and I_Ks_ currents were recorded at 37 °C from isolated hiPSC-CMs plated on glass coverslips as described in detail in the Supplemental Methods section. Action potentials were recorded in perforated patch mode under 1 Hz pacing.

### MultiElectrode Arrays

HiPSC-CMs were plated on 24-well multiwell MEAs (MultiChannel Systems) coated with bovine fibronectin (Merck) as previously described ^43,44^. The recordings were performed on spontaneously beating hiPSC-CMs at 37°C. The detailed description of drug treatments and analysis of the recordings is present in the Supplemental Methods section.

### Pro-arrhythmic compound selection

The selection of pro-arrhythmic compounds was performed using the CredibleMeds® QT prolonging drugs database ^45^ intersected with OpenFDA database (https://open.fda.gov/). OpenFDA data were used to prioritize compounds based on their occurrence in the FDA Adverse Event Reporting System. The following query terms were utilized to extract the top 100 compounds from the OpenFDA database: “*Cardiac failure*”, “*Torsade de pointes*”, “*Electrocardiogram QT prolonged*”, “*Cardiac arrest*”, “*Ventricular extrasystoles*”, “*Ventricular arrhythmia*”. The common compounds between databases were 46 (Supplemental Table 3). We excluded compounds that were unlikely to provide pro-arrhythmic effects in a pure culture of hiPSC-CMs (such as diuretics or drugs associated with chronic cardiotoxicity) as beyond the scope of this study. Seven drugs were then selected in the highest-risk category, i.e. those contraindicated in congenital LQTS and are known to pose a significant risk for developing drug-induced Torsades de Pointes (TdP). Detailed information on the compounds is provided in Supplemental Table 4.

### Data Analysis and Statistics

RStudio (version 2023.09.1) and R (version 4.3.1) were used for data analysis. Multiple group comparisons were conducted using the Kruskal-Wallis test, followed by Dunn’s test with Benjamini-Hochberg correction. Pairwise comparisons were performed using the Wilcoxon test. Categorical variable comparisons were assessed with Fisher’s exact test. A statistical significance level of p ≤ 0.05 was used for all tests. Figures denote significance as *p ≤ 0.05, **p ≤ 0.01, and ***p ≤ 0.001, while exact p-values are reported in text and tables. Numerical data are reported as mean ± standard error of the mean (SEM), and plots display mean ± SEM, combined with scattered individual data points when relevant. Categorical data are presented as bar plots.

### Machine learning model for variant risk prediction

A random forest dichotomous classification model was implemented using the comprehensive *tidymodels* framework ^46^. The dataset was stochastically divided into training and validation subsets with a ratio of 70:30, ensuring that drug readouts were evenly distributed between the two datasets. A v-fold (v = 10) cross validation was performed to evaluate the model performance. To further evaluate the classification performance of the model for each genetic variant, a leave-one-out cross validation was performed by recursively removing the data for each variant. The accuracy of variant prediction was calculated as the percentage of correctly predicted readouts (rows) on the total number of readouts of the dataset. Overall score of variant prediction was calculated as a sum of accuracies of each variant risk classification per number of variants. The datasets, models and R code used for the analyses are available in the lab’s GitHub repository: https://github.com/invitroheart.

## Results

### Patient cohort characteristics

The study included ten patients, with their clinical information summarized in Table 1. The study cohort comprised a LQT1 family trio (Supplemental Figure 3), which includes an asymptomatic mother carrying the *KCNQ1* p.R190W variant, an asymptomatic father carrying the *KCNQ1* p.R594Q variant, and their daughter affected by Jervell and Lange-Nielsen syndrome^47^ (JLNS) carrying both variants in compound heterozygosity ^48^. Four patients from a large South African family carrying the *KCNQ1* p.A341V variant associated with a very severe LQT1 phenotype ^49–51^; two of these patients were asymptomatic while the other two were severely symptomatic, carrying polymorphisms in the modifier gene *NOS1AP* (one patient in heterozygous and one patient in homozygous state) associated with a prolonged QT interval and with an increased risk of life-threatening events ^52^. Among the LQT2 cases, two patients carried the *KCNH2* p.R366X variant: one symptomatic and one asymptomatic. Both were treated with beta-blockers. The symptomatic patient also underwent left cardiac sympathetic denervation due to difficulties in optimizing beta-blocker therapy because of asthenia. No recurrence of symptoms was observed on therapy. A third symptomatic LQT2 patient, carrying the de novo *KCNH2* p.A561V variant, experienced cardiac arrests and was treated with beta-blockers and an ICD implantation.

### Genetic variant risk level assignment

To define the threshold for categorizing variants associated with different risks of cardiac events (Low Risk vs High Risk) we analyzed the frequency distribution of cardiac events across reported P/LP genetic variants in *KCNQ1* and *KCNH2* genes (1099 patients carrying 162 variants in *KCNQ1* and 623 patients carrying 175 variants in *KCNH2*) (Figure 1, A). For both genes, two distinct peaks in the distribution were identified: one at 0% frequency (variants with no reported cardiac events) and another at ≥25% frequency (variants where at least 25% of carriers experienced cardiac events). Based on this analysis, a threshold of 25% was selected to differentiate these populations. Using this threshold, the variants in our study were categorized as follows: High Risk (*KCNQ1* p.R594Q and *KCNQ1* p.R190W & p.R594Q, *KCNQ1* p.A341V, and *KCNH2* p.A561V) and Low Risk (WT, *KCNQ1* p.R190W, and *KCNH2* p.R366X) (Figure 1, B).

**Figure 1.**
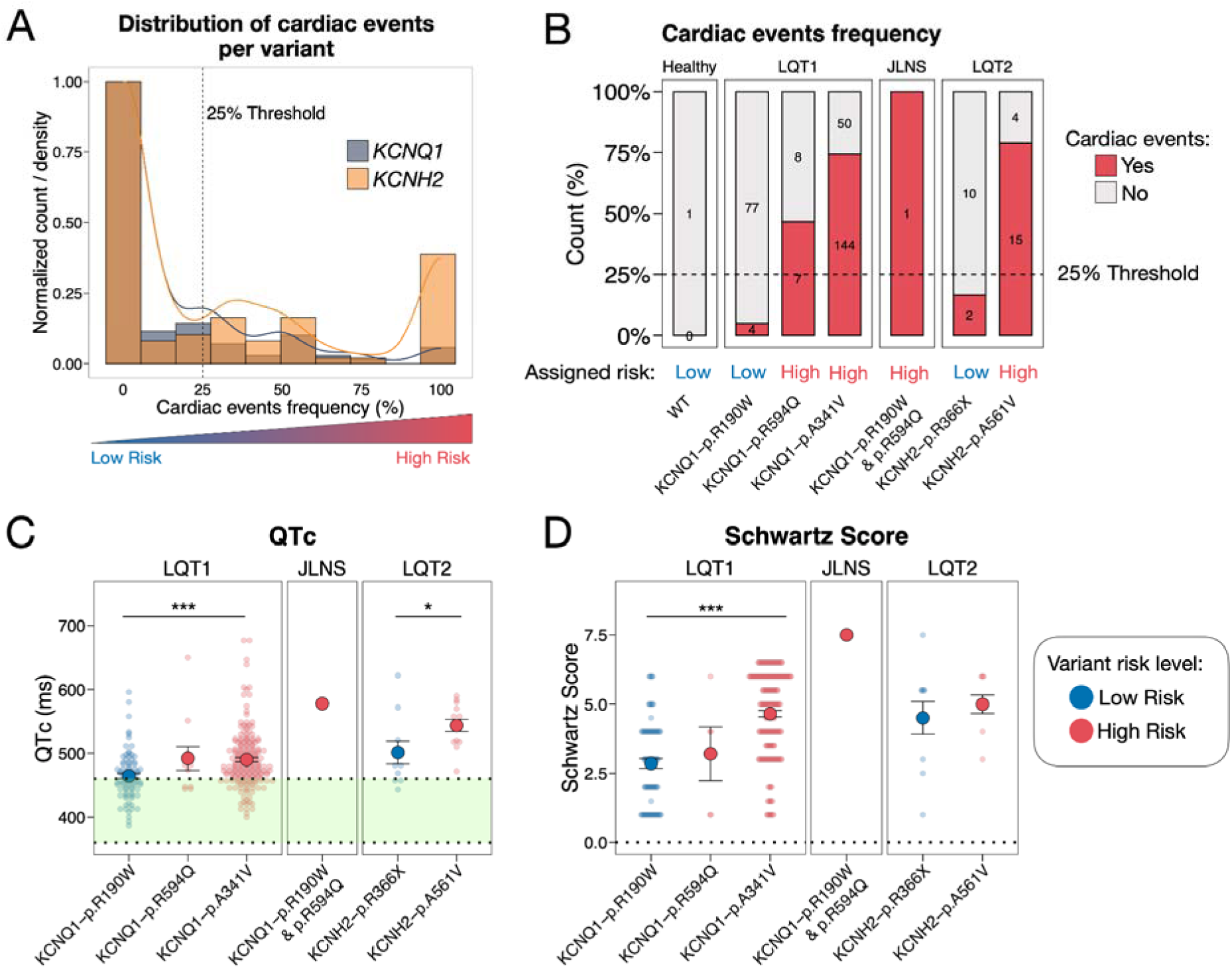
Clinical parameters and variant risk level assignment. A. Frequency of cardiac events across pathogenic/likely pathogenic genetic variants in *KCNQ1* (blue) and *KCNH2* (orange) genes. B. Percentage of patients with cardiac events for each variant, with assigned variant risk level indicated. C, D. Baseline QTc and Schwartz score values for each variant (N = 1–183).

Consistent with the incidence of cardiac events, carriers of High Risk variants *KCNQ1* p.A341V and *KCNH2* p.A561V had significantly longer QTc values compared to carriers of the Low Risk variants *KCNQ1* p.R190W and *KCNQ1* p.R366X respectively (Figure 1, C, Supplemental Table 5). In addition, the Schwartz scores of *KCNQ1* p.A341V carriers were higher than those of *KCNQ1* p.R190W (Figure 1, C, Supplemental Table 5). Notably, the *KCNQ1* p.R190W & p.R594Q variant, observed in a single carrier, presented with deafness and exhibited markedly prolonged QTc (578 ms) and elevated Schwartz score (7.5), consistent with the severe JLNS phenotype.

### *In vitro* characterisation of genetic variants selected for the study

Four out of six variants had functional characterization from literature: *KCNQ1* p.R594Q, *KCNQ1* p.A341V, *KCNH2* p.R366X, *KCNH2* p.A561V leading to loss-of-function, with *KCNQ1* p.R594Q and *KCNH2* p.A561V being also trafficking-deficient (Supplemental Table 6). We complemented the characterization providing data for the hiPSC-CMs derived from a family trio (*KCNQ1* p.R190W, *KCNQ1* p.R594Q, and *KCNQ1* p.R190W & p.R594Q) (Figure 2).

**Figure 2.**
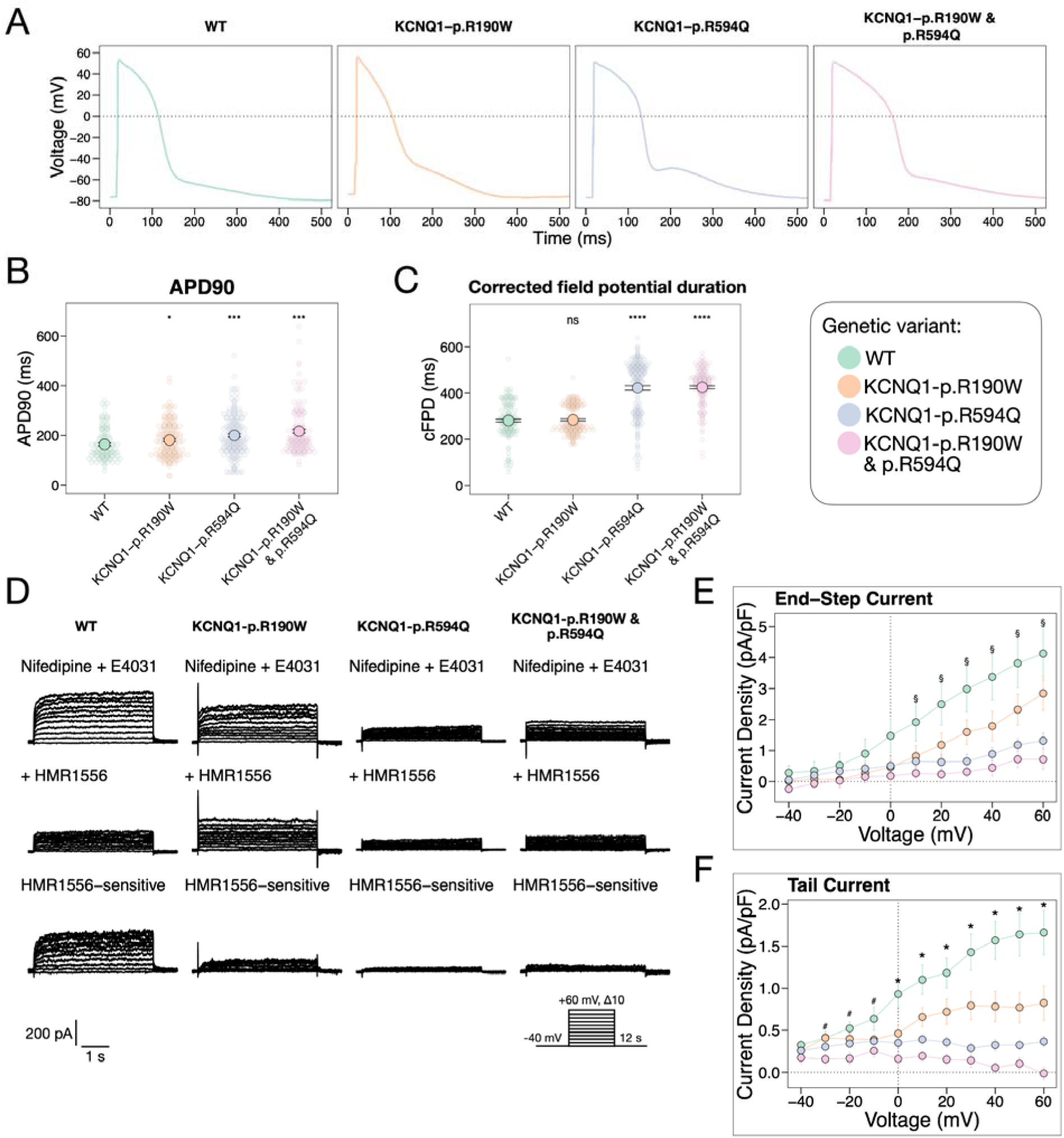
Characterization of hiPSC-CMs carrying genetic variants *KCNQ1* p.R190W and *KCNQ1* p.R594Q. A. Representative action potential traces for each hiPSC-CM variant, elicited at 1 Hz stimulation. B. Action potential duration at 90% repolarization (APD90). C. Corrected field potential duration (cFPD) measurements in hiPSC-CMs for each variant (N = 157–180). D. Representative I_Ks_ current traces (lower panel), obtained by subtracting HMR1556-insensitive current (middle panel) from the total inward potassium current recording (upper panel). E, F End-step and tail I_Ks_ current-voltage (I-V) relationships. * denotes p < 0.05 for all groups vs WT, # for WT vs *KCNQ1* p.R190W & p.R594Q, § for WT vs KCNQ1 p.R594Q and vs KCNQ1 p.R190W & p.R594Q.

We observed the repolarization prolongation at single-cell level (APD90) and a reduced ability to follow high-frequency pacing in isolated hiPSC-CMs carrying *KCNQ1* p.R190W, *KCNQ1* p.R594Q, and *KCNQ1* p.R190W & p.R594Q compared to WT (Figure 2, A, B; Supplemental Figure 4, A, B). HiPSC-CM monolayers carrying *KCNQ1* p.R594Q and *KCNQ1* p.R190W & p.R594Q demonstrated also corrected FPD prolongation in comparison to WT (422.2 ± 126.7 ms and 424.9 ± 78.7 ms vs 281.8 ± 77.4 ms respectively, p ≤ 0.001) (Figure 2, C). Measurement of I_Ks_ with patch clamp demonstrated decreased end-step and tail current densities in hiPSC-CMs carrying *KCNQ1* p.R190W, compared to WT, and almost absent I_Ks_ in hiPSC-CMs carrying *KCNQ1* p.R594Q and *KCNQ1* p.R190W & p.R594Q, in line with their APD prolongation (Figure 2, D-F).

### Baseline electrophysiology of hiPSC-CM cohort

Baseline hiPSC-CM electrophysiology assessed with MEA measurements accurately reflected the risk level assigned to each variant. Specifically, hiPSC-CMs from the High Risk group exhibited increased baseline FPD, corrected FPD (cFPD), and RR interval duration (Figure 3, A-D; Table 2) in comparison to wild type hiPSC-CMs from the Low Risk group. The propensity for irregular beating patterns at baseline was significantly higher in the High Risk group (20.2% of wells in High Risk group vs 8.7% in Low Risk group, p ≤ 0.001) (Figure 3, E).

**Figure 3.**
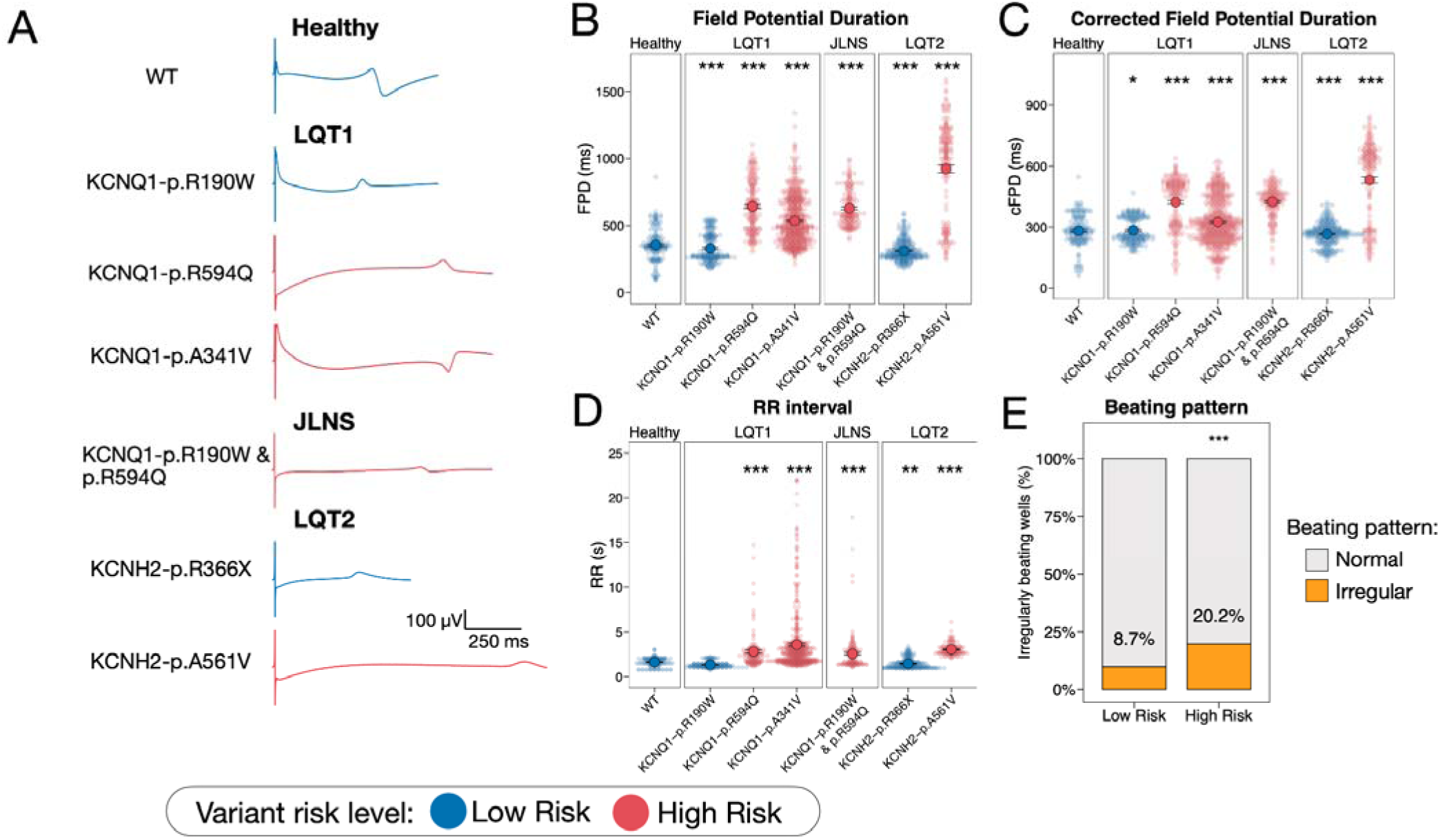
Baseline electrophysiology of hiPSC-CMs carrying High Risk and Low Risk genetic variants. A. Representative field potentials obtained for each variant. B-D. Baseline field potential duration (FPD), corrected FPD (cFPD), and beat-to-beat interval (RR) measurements of hiPSC-CMs for each variant; asterisks indicate statistical significance with the WT group. N = 154-547. E. Incidence of regular (gray) or irregular (yellow) beating patterns for hiPSC-CMs carrying Low Risk and High Risk variants (N = 589-1038).

**Table 2.** Baseline electrophysiological parameters obtained from MEA recording for each variant.

| Variant | N | FPD, ms | cFPD, ms | RR, s |
| --- | --- | --- | --- | --- |
| WT | 154 | 356 ± 120.3 | 281.8 ± 77.4 | 1.63 ± 0.49 |
| KCNQ1-p.R190W | 180 | 331.3 ± 104.9, p = 0.07 | 284.6 ± 60.6, p = 0.69 | 1.33 ± 0.34, p ≤ 0.001 |
| KCNQ1-p.R594Q | 173 | 643.7 ± 190.2, p ≤ 0.001 | 422.2 ± 126.7, p ≤ 0.001 | 2.82 ± 2.6, p ≤ 0.001 |
| KCNQ1-p.A341V | 547 | 536.9 ± 173.2, p ≤ 0.001 | 325.1 ± 109.2, p ≤ 0.001 | 3.57 ± 3.71, p ≤ 0.001 |
| KCNQ1-p.R190W & p.R594Q | 157 | 629.8 ± 133, p ≤ 0.001 | 424.9 ± 78.7, p ≤ 0.001 | 2.56 ± 2.05, p ≤ 0.001 |
| KCNH2-p.R366X | 255 | 311.4 ± 74.3, p ≤ 0.001 | 265.9 ± 53.4, p = 0.06 | 1.45 ± 0.56, p ≤ 0.001 |
| KCNH2-p.A561V | 161 | 923.1 ± 355.4, p ≤ 0.001 | 531.1 ± 197.3, p ≤ 0.001 | 3.04 ± 0.61, p ≤ 0.001 |

### HiPSC-CMs from High Risk and Low Risk groups demonstrate distinct drug responses to ion channel blockers

To further discriminate hiPSC-CMs carrying variants with different risk levels we obtained concentration-response curves for 11 selected drugs (Figure 4, 5; Supplemental Figures 5-16; Supplemental Table 7). Selective ion channel blockers E4031 (I_Kr_, Kv11.1), HMR-1556 (I_Ks_, Kv7.1), nifedipine (I_CaL_, CaV 1.2), and tetrodotoxin (I_Na_, Nav 1.5) were used to assess ion channel-specific drug responses as previously proposed by multiple ion channel effects (MICE) approach for drug testing assays ^53^. Since I_Ks_ blockers were previously reported to be more effective with reduced cardiac repolarization reserve caused by *KCNH2* genetic variants or I_Kr_ pharmacological block ^54^, we also tested the effect of HMR1556 on hiPSC-CMs pre-treated with 20 nM I_Kr_ blocker E4031.

**Figure 4:**
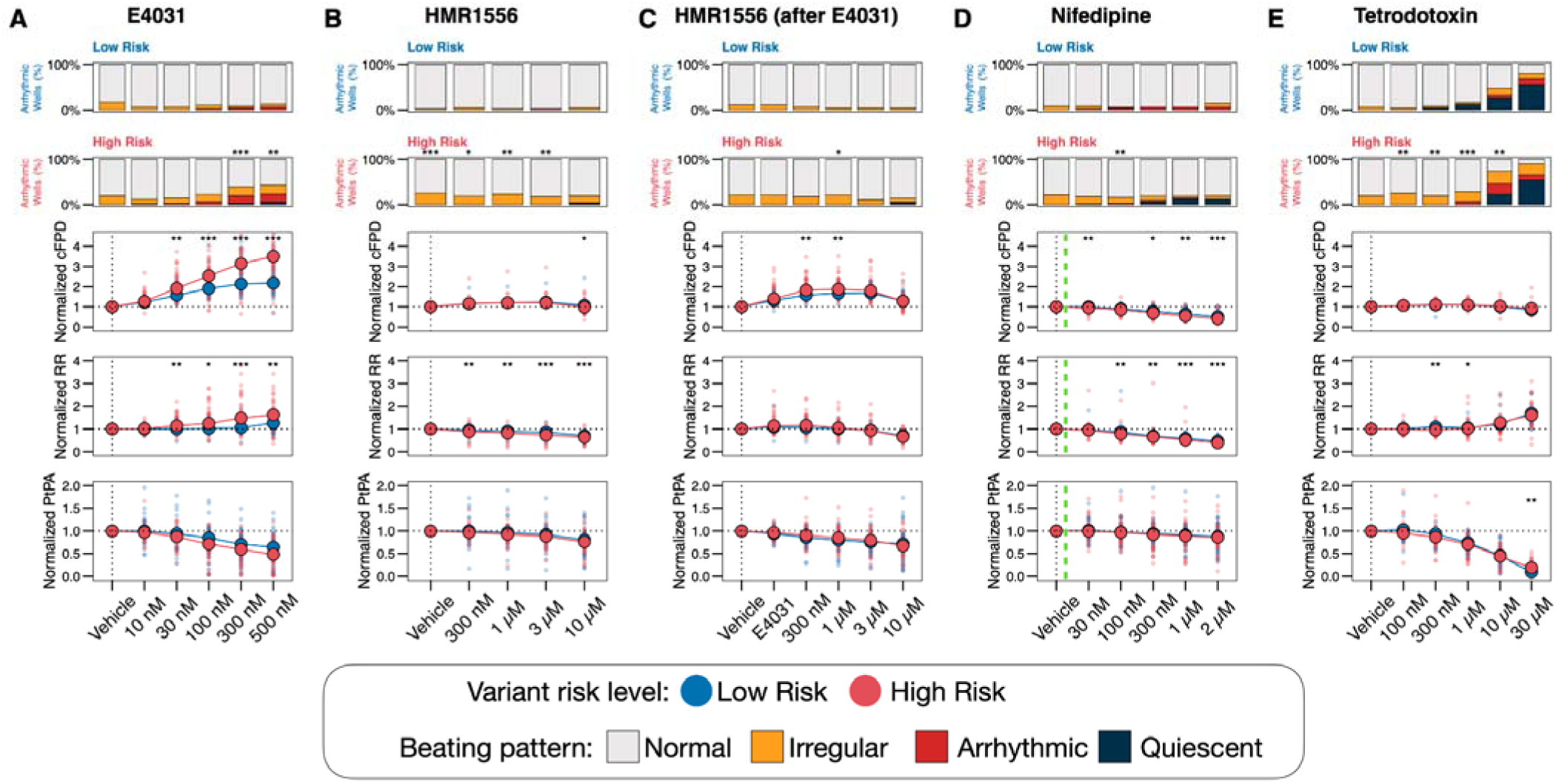
HiPSC-CMs from Low Risk and High Risk groups exhibit differential responses to ion channel blockers. A-E: Concentration-dependent response curves to ion channel blockers E4031, HMR1556, HMR1556 following E4031 pre-treatment, nifedipine, and tetrodotoxin. Responses are shown for normalized corrected field potential duration (normalized cFPD), normalized RR interval (normalized RR), and normalized peak-to-peak amplitude (normalized PtPA). Bar plots illustrate the distribution of beating pattern abnormalities. Green dashed lines indicate reported Cmax values for each drug. Asterisks indicate statistical significance of comparisons between the High Risk and Low Risk groups. N = 19-80 per group per drug concentration.

Among pure ion channel blockers, we observed a differential response to the I_Kr_ blocker E4031 and to I_Ks_ blocker HMR1556 after E4031 pretreatment between hiPSC-CMs carrying High Risk and Low Risk variants. Specifically, hiPSC-CMs with High Risk variants showed a greater concentration-dependent prolongation of cFPD following treatment with E4031 (up to 350.0 ± 183.2% vs Vehicle for High Risk and up to 218.2 ± 69.9% vs Vehicle for Low Risk, p ≤ 0.001). Furthermore, E4031 increased the RR interval duration in hiPSC-CMs carrying High Risk variants more than in those carrying Low Risk variants (Figure 4, A). The High Risk group also demonstrated a higher incidence of beating abnormalities at high concentrations of E4031 (Figure 4, A). HMR1556 treatment alone resulted in a small cFPD prolongation that was similar between the High Risk and Low Risk groups (Figure 4, B). In contrast the combination of E4031 pre-treatment followed by HMR1556 concentration-response led to a significant cFPD prolongation (up to 188.4 ± 40.6% vs. Vehicle for High Risk and up to 169.2 ± 26.4% vs. Vehicle for Low Risk, p = 0.004) (Figure 4, C). Treatment with the I_CaL_ blocker nifedipine led to a cFPD and RR interval decrease (Figure 4, D). High concentrations of the I_Na_ blocker tetrodotoxin (10 µM and 30 µM) led to a decrease in peak-to-peak amplitude and prolongation of the RR interval, along with an increased incidence of beating abnormalities (Figure 4, E). The High Risk group exhibited significantly more beating abnormalities at 10 µM of tetrodotoxin compared to the Low Risk group (73.6% vs. 46.9%, respectively; p = 0.003).

### HiPSC-CMs carrying High Risk and Low Risk P/LP variants demonstrate distinct drug responses to proarrhythmic drugs

In addition to pure ion channel blockers seven proarrhythmic compounds were selected using the OpenFDA and CredibleMeds databases, as described in the Methods: β2-adrenergic receptor agonist albuterol (salbutamol); the antibiotics clarithromycin, ciprofloxacin, and moxifloxacin; the antipsychotic drugs chlorpromazine and haloperidol; the antiarrhythmic drug dofetilide. Among these compounds dofetilide, haloperidol, and the β2-adrenergic receptor agonist salbutamol demonstrated differential concentration-response curves between High Risk and Low Risk groups (Figure 5). Specifically, responses to the antibiotics clarithromycin and ciprofloxacin were similar between the two groups (Figure 5 A, C). The highest concentration of clarithromycin tested (10 µM) led to arrhythmias and cessation of beating (Figure 5 A). Another antibiotic, moxifloxacin, demonstrated a similar effect, with notable toxicity at a concentration of 100 µM, expressed by cFPD and RR increases, a drop in peak-to-peak amplitude, arrhythmias, and cessation of beating similar for both groups (Figure 5, F). Treatment of hiPSC-CMs with the antipsychotic drug chlorpromazine resulted in moderate cFPD prolongation that was similar between the two groups (Figure 5, B). hiPSC-CMs carrying High Risk variants showed a concentration-dependent increase in cFPD following treatment with the dofetilide (up to 317.5 ± 128.5% vs. Vehicle for High Risk and up to 208.2 ± 61.4% vs. Vehicle for Low Risk, p ≤ 0.001) (Figure 5, D) and haloperidol (up to 253.1 ± 134.0% vs. Vehicle for High Risk and up to 178.2 ± 50.6% vs. Vehicle for Low Risk, p ≤ 0.001) (Figure 5, E). Salbutamol decreased the RR interval of hiPSC-CMs carrying High Risk variants to a greater extent compared to those carrying Low Risk variants (Figure 5, G).

**Figure 5.**
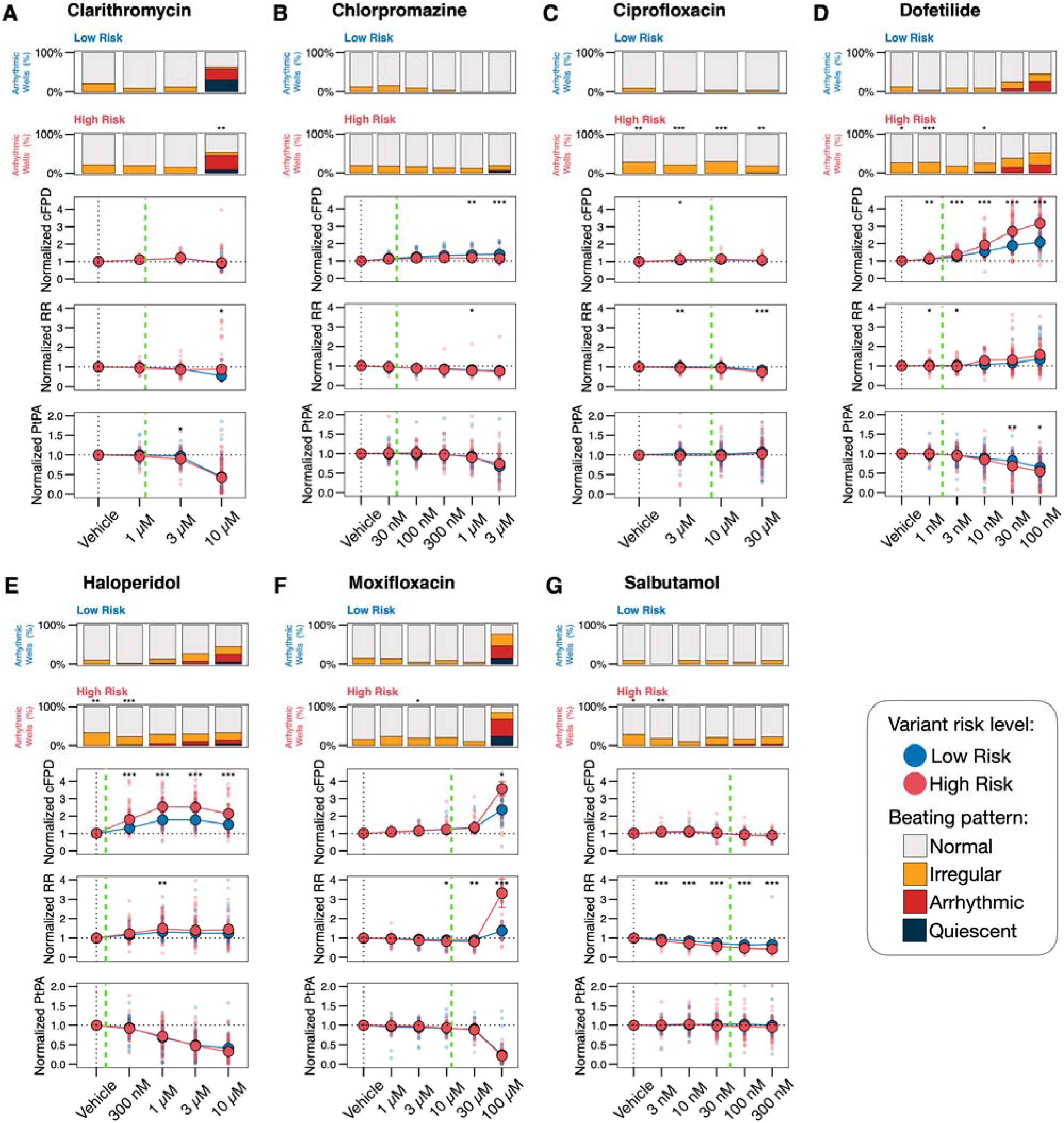
HiPSC-CMs from Low Risk and High Risk groups exhibit differential responses to proarrhythmic drugs. A-G: Concentration-dependent response curves for proarrhythmic drugs clarithromycin, chlorpromazine, ciprofloxacin, dofetilide, haloperidol, moxifloxacin, and salbutamol. Responses are presented for normalized corrected field potential duration (normalized cFPD), normalized RR interval (normalized RR), and normalized peak-to-peak amplitude (normalized PtPA). Bar plots illustrate the distribution of beating pattern abnormalities. Green dashed lines indicate reported Cmax values for each drug. Asterisks indicate statistical significance of comparisons between the High Risk and Low Risk groups. N = 26-121 per group per drug concentration.

### Gene-specific drug response differences between in hiPSC-CMs carrying High Risk and Low Risk P/LP variants

To evaluate gene-specific drug responses, we compared the concentration-response curves between High and Low Risk P/LP variants, separating hiPSC-CMs with *KCNQ1* variants (LQT1, JLNS) from those with *KCNH2* variants (LQT2). We analyzed responses to I_Kr_-blocking drugs – dofetilide, E4031, and haloperidol – which demonstrated the strongest discrimination between High Risk and Low Risk P/LP variants.

Gene-specific features emerged in drug responses between hiPSC-CMs with High and Low Risk P/LP variants. High Risk hiPSC-CMs carrying *KCNQ1* variants showed significant FPD prolongation upon drug treatment (Figure 6A–C; Supplemental Table 8), while, in contrast, High Risk *KCNH2* variant carriers demonstrated cFPD prolongation similar to the Low Risk *KCNH2* group, but exhibited a marked decrease in peak-to-peak amplitude and an increased incidence of abnormal beating patterns (Figure 6D–F; Supplemental Table 9).

**Figure 6.**
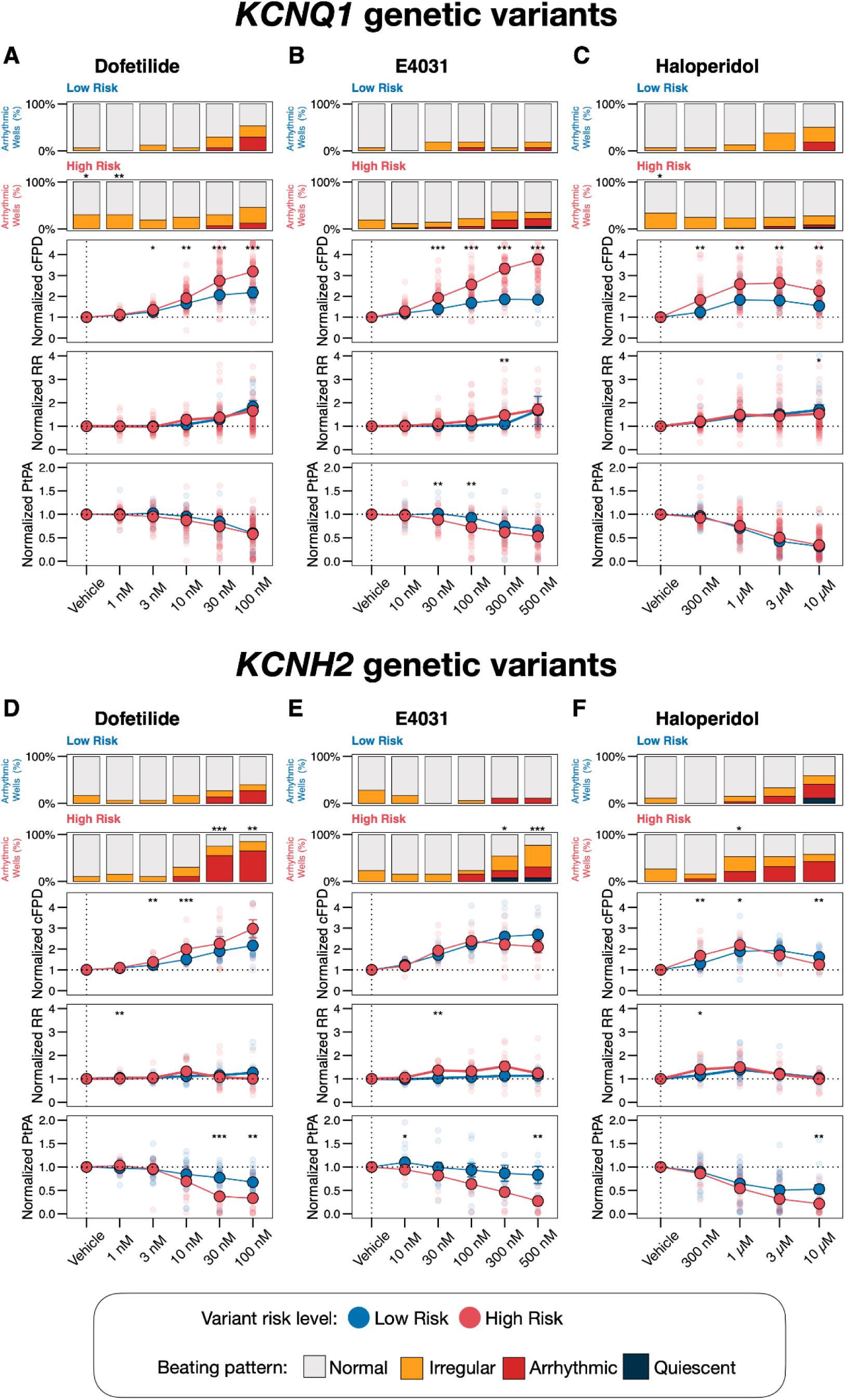
HiPSC-CMs carrying High Risk and Low Risk variants demonstrate gene-specific drug responses. A-F. Concentration-dependent response curves for dofetilide, E4031, and haloperidol on normalized corrected field potential duration (normalized cFPD), normalized RR interval (normalized RR), and normalized peak-to-peak amplitude (normalized PtPA). Bar plots illustrate the distribution of beating pattern abnormalities. Asterisks indicate statistical significance of comparisons between High Risk and Low Risk groups. N = 8-101 per group per drug concentration.

### Machine learning model stratifies P/LP variant risk from hiPSC-CM multielectrode array data

We curated a MEA dataset from eleven patient-specific hiPSC lines carrying seven variants, exposed to a panel of 11 drugs. The complete dataset comprises 10,410 data points and it was used to train a machine learning classifier for stratifying P/LP variant risk based on in vitro drug responses in patient-specific hiPSC-CMs. To ensure robust and unbiased predictions, we deliberately included all drug-responses, even those from drugs that did not show apparent discrimination between High Risk and Low Risk variants according to concentration-response curves. The random forest model demonstrated robust classification performance with 89% accuracy and excellent discriminative ability, with an area under the receiver operating characteristic (ROC) curve (AUC) of 96% (Figure 7, A). The model also demonstrated a strong discriminative ability for LQTS subtype, achieving 79% classification accuracy and an AUC of 94% (Figure 7, B).

**Figure 7.**
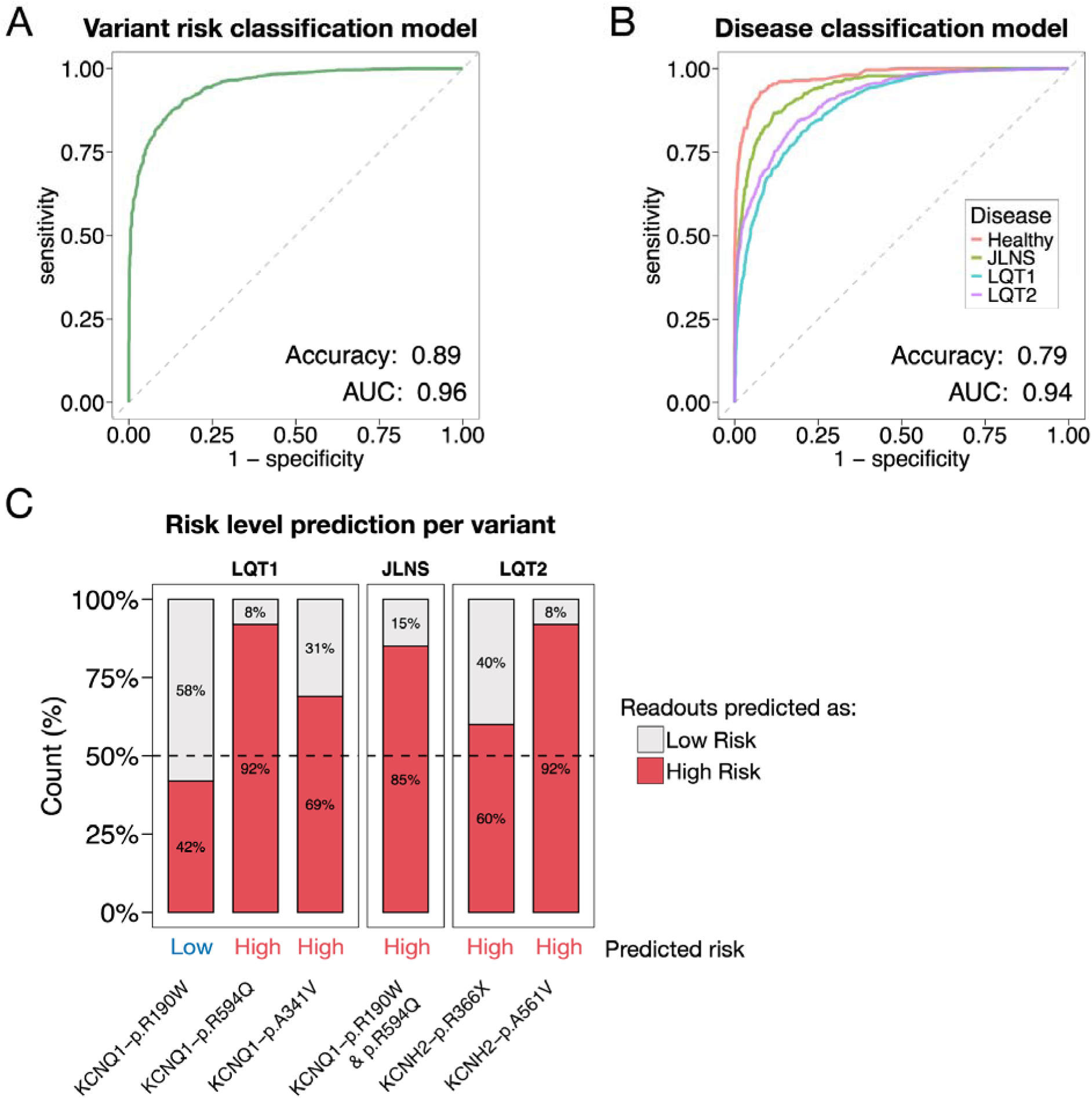
Variant risk levels and disease type prediction using MEA measurements and machine learning model. A. Receiver Operating Characteristic (ROC) curve illustrating the performance of a classification model for variant risk prediction. B. ROC curves representing the model’s performance in predicting disease type. C. Model validation for single-variant risk level predictions. The proportion of readouts predicted as High Risk (red) and Low Risk (gray) is shown for each variant. The 50% threshold line delineates variants classified as High Risk (above) from those classified as Low Risk (below).

### Machine learning validation confirmed model accuracy for predicting individual variant risk

To further evaluate the model’s generalization performance and its ability to predict risk levels for novel variants, we performed leave-one-out cross-validation by sequentially excluding each variant from the training dataset and predicting its risk level as if it were a novel variant. The model correctly classified 6 out of 7 variants into their appropriate risk categories, with one exception (*KCNH2* p.R366X), achieving an overall prediction accuracy of 72.6% (Figure 7, C, Table 3).

**Table 3.** LQTS variant risk level prediction accuracy for each genetic variant included in the study.

| **Variant** | **Disease** | **Variant risk level** | **Predicted variant risk level** | **Accuracy** |
| --- | --- | --- | --- | --- |
| KCNQ1-p.R190W | LQT1 | Low Risk | Low Risk | 0.58 |
| KCNQ1-p.R594Q | LQT1 | High Risk | High Risk | 0.92 |
| KCNQ1-p.A341V | LQT1 | High Risk | High Risk | 0.69 |
| KCNQ1-p.R190W & p.R594Q | JLNS | High Risk | High Risk | 0.85 |
| KCNH2-p.R366X | LQT2 | Low Risk | High Risk | 0.4 |
| KCNH2-p.A561V | LQT2 | High Risk | High Risk | 0.92 |
| **Overall accuracy of variant prediction 0.73** |  |  |  |  |

Accuracy above 0.5 indicates correct variant risk level prediction.

## Discussion

Risk stratification for LQTS patients remains a significant clinical challenge, particularly for carriers of ultra-rare P/LP variants or for those identified in a single proband. These shortcomings impact our ability to identify patients who are at low or high risk of developing cardiac events that could potentially lead to SCD. Here, we developed a machine learning pipeline to classify P/LP variant risk based on *in vitro* electrophysiological readouts from patient-specific hiPSC-CM lines carrying genetic variants associated with varying frequencies of cardiac events (Low Risk or High Risk).

Our findings support prior evidence demonstrating that hiPSC-CMs can effectively recapitulate the clinical phenotype of LQTS ^20–24^. This was particularly evident in hiPSC-CMs carrying severe genetic variants, such as compound heterozygous *KCNQ1* p.R190W & p.R594Q, *KCNQ1* p.A341V, and *KCNH2* p.A561V, where a clear prolongation in the repolarization duration was clearly observed *in vitro*.

We further strengthened the dataset beyond baseline measurements by challenging each variant with ion channel blockers and pro-arrhythmic compounds, thus creating variant-specific drug response profiles not immediately obtainable in clinical practice. This is supported by previous research demonstrating excellent capacity of hiPSC-CMs to discriminate high/low responders to an acute sotalol administration ^55^ or to tyrosine kinase inhibitors ^56,57^. To overcome the high baseline variability typical of hiPSC-CM measurements, often observed in cohort studies, we used relative readouts obtained from normalized drug responses to provide a more accurate discrimination between disease phenotypes. Indeed, High Risk variants were clearly discriminated from their exacerbated response to certain pro-arrhythmic drug treatments, leading to more severe changes in FPD, cFPD, and RR intervals or to a higher occurrence of beating pattern abnormalities.

The panel of pure ion channel blockers targeting four key ion channels responsible for action potential regulation revealed that pharmacological I_Kr_ blockade alone already provides effective variant risk discrimination. Conversely, treatment of hiPSC-CMs with the I_Ks_ blocker HMR1556 caused only minimal FPD prolongation ^58,59^. Reducing the repolarization reserve by pre-treating hiPSC-CMs with I_Kr_ blocker E4031 ^54,60^ increased the sensitivity to HMR1556 and discriminated between High Risk and Low Risk groups. As expected, the I_CaL_ blocker nifedipine and the I_Na_ blocker tetrodotoxin were not effective in distinguishing High Risk from Low Risk groups within our LQT1 and LQT2 cohort, which is expected since no pathogenic variants were identified in genes encoding for calcium and sodium channel subunits. These blockers could be useful for identifying Ca² [or Na [handling abnormalities to other LQTS subtypes beyond LQT1 and LQT2, e.g. LQT3 or CALM-LQTS ^22,61^.

Among the pro-arrhythmic drugs selected based on analyses of the OpenFDA and CredibleMeds databases, acute treatment of hiPSC-CMs with the potent I_Kr_ blocker dofetilide and the antipsychotic drug haloperidol demonstrated clear concentration-dependent cFPD prolongation, consistently distinguishing between the High Risk and Low Risk groups. Acute treatment with salbutamol, a β_2_-adrenergic agonist commonly used as a bronchodilator and typically precluded to LQTS patients ^62^, differentially increased the spontaneous beating frequency in hiPSC-CMs; this allowed the discrimination of High Risk variants, which revealed a higher sensitivity, from Low Risk variants and could be influenced by a difference in beating frequencies at baseline between these two groups. No arrhythmic events were observed supporting previous studies on commercially available hiPSC-CMs ^63^. Treatment of hiPSC-CMs with ciprofloxacin, clarithromycin, and moxifloxacin within therapeutic concentration ranges resulted in moderate FPD prolongation as similarly emerged in commercially available wild-type hiPSC-CMs ^31,64^.

There is limited information available on gene-specific drug responses in LQT1 and LQT2 both *in vivo* and *in vitro* ^65–67^. In our study, three drugs with increased potency to block hERG (E4031, dofetilide, and haloperidol) well discriminated diseased hiPSC-CMs carrying High Risk variants in either *KCNQ1* or *KCNH2* genes. This observation aligns with findings from adult transgenic rabbit models, where LQT2 rabbits exhibited greater susceptibility to drug-induced TdP than LQT1 rabbits after administration of the anesthetic propofol, with a known hERG channel blocking effect^66^. These results are consistent with the concept of a severely compromised repolarization reserve in LQT2 cardiomyocytes, which renders them more vulnerable to proarrhythmic triggers ^60^.

Variant risk prediction based on multiparametric readouts is a novel and rapidly developing field. A recent study combined clinical data, trafficking assays and automated patch clamp data from heterologous expression systems, to score the risk of *KCNH2* variants associated with LQT2 ^68^. The use of hiPSC-CMs provides enhanced precision by addressing the limitations of heterologous systems that lack cardiac-specific context. Having demonstrated that hiPSC-CM drug responses effectively discriminate between High Risk and Low Risk P/LP variants, we developed the ML model trained on *in vitro* hiPSC-CM electrophysiological readouts able to predict retrospectively severity of variants with high precision – a methodology that, to our knowledge, has not been previously applied to LQTS. While we deliberately chose well-known P/LP genetic variants with a loss-of-function mechanism for our study, the model’s versatility extends beyond retrospective analyses to the classification of ultra-rare variants and VUS. Importantly, the ML model trained for disease discrimination predicted each disease subtype with a 79% accuracy. This robust performance indicates that disease-specific electrophysiological features are distinct from variant risk classification features, providing solid evidence to expand the model incorporating other LQTS subtypes.

Successful application of the model to predict the risk level of single variants on which it was not originally trained represents a high-level validation, further demonstrating applicability of our approach for assessing novel, previously unreported variants. The observed discrepancy in predicting the risk level for the *KCNH2* p.R366X variant underscores the impact of patient-specific drug responses. Although assigned as Low Risk due to a low cardiac event frequency (16.7%), two hiPSC-CM lines from different LQT2 patients exhibited divergent drug response patterns. One line, derived from a patient with no cardiac events, aligned with Low Risk variants, whereas the other, from a patient with documented cardiac events, markedly prolonged QTc (622 ms), and LSCD performed, with those of High Risk. This highlights the importance of separating the genetic background from the primary disease-causing variant, particularly for P/LP variants that fall within the intermediate range of the risk level distribution. Indeed, in the case of High Risk *KCNQ1* p.A341V variant, despite differences in genetic background (such as *NOS1AP* genetic variants) the primary variant itself is so severe that the probability of misclassification is lower. These findings emphasize the necessity of integrating data from multiple sources for accurate variant risk classification, including multicentral *in vitro* data, clinical cohort records, structural variant pathogenicity predictions, and Bayesian penetrance estimates ^37^.

In conclusion, we established a robust method for stratifying P/LP variant risk by integrating ML classification to electrophysiological recordings in patient-specific hiPSC-CMs. This approach enhances the risk stratification of LQTS variants beyond what is currently possible from clinical data alone, providing value for variants with limited clinical information.

## Study limitations

First, experiments were performed on hiPSC-CMs which, despite significant progress in maturation techniques, still exhibit an immature phenotype compared to adult cardiomyocytes ^69,70^. This immaturity could partially explain blunted or exacerbated drug responses for specific compounds.

Secondly, we focused on six LQTS-causing LP/P variants. While this demonstrates proof of principle for retrospective variant risk stratification, further investigations are necessary to extend this approach to a broader range of variants or LQTS subtypes.

Third, while our model simplifies clinical risk into a binary classification, further evolutions of this approach could consider incorporating multiple classification states to accommodate intermediate disease phenotypes based on both qualitative and quantitative cardiac events associated with each variant.

## Data Availability

The datasets, models and R code used for the analyses are available in the lab GitHub repository.

## Abbreviations and acronyms

APD: Action Potential Duration
FPD: Field Potential Duration
iPSC-CMs: Cardiomyocytes derived from human induced pluripotent stem cells
JLNS: Jervell and Lange-Nielsen Syndrome
LQTS: Long QT Syndrome
MEA: Multielectrode Arrays
ML: Machine Learning
P/LP: Pathogenic/Likely Pathogenic
TdP: Torsades de Pointes
QTc: QT interval corrected for heart rate
WT: Wild type

