## Supplemental Information for "Genetic variants risk assessment for Long QT Syndrome through machine learning and multielectrode array recordings"

### **Supplemental Appendix**

### **Supplemental methods**

A list of main reagents used in the study is reported in Supplemental Table 2.

**HiPSC cardiac differentiation**

Cardiac differentiation was started 2-4 days after hiPSC plating, when hiPSCs reached 70-90% confluency. At this stage (Day 0), the culture medium was switched to RBins- medium, which consists of RPMI1640 supplemented with 1% of B27 supplement without insulin (Gibco), and 6 µM CHIR99021 (Selleckchem) was added to induce the differentiation. For the next two days, fresh RBins- medium was added (⅔ of the initial volume on day 1 and ⅓ on day 2). On day 3, the medium was switched to RBins- medium supplemented with 5 µM IWR1 (Merck). After two days of treatment with IWR1, the medium was replaced with RBK medium, consisting of RPMI1640 supplemented with 1% B27 Supplement (Gibco) and 1% Knock-Out Serum Replacement (Gibco). Starting from day 7 hiPSC-CMs were purified through glucose starvation ^1^ and cryopreserved in Bambanker (Nippon Genetics) at days 9-16. For the subsequent experiments hiPSC-CMs were thawed, replated at low density and expanded as previously reported ^2^. Data were collected from at least three independent differentiations of each hiPSC line for each experiment.

#### **Patch-clamp current-clamp and voltage-clamp recordings**

Expanded hiPSC-CMs were dissociated with TrypLE Select Enzyme (10X) (Gibco) and plated in monolayers at a density 3 * 10^5^/cm^2^ on 12-well plates coated with Matrigel (Corning) in RBK medium. The next day, RBK was replaced with a lipid-rich maturation medium (MM) ^3^. Metabolic maturation was performed for two weeks and MM was refreshed twice a week. For action potential (AP) recordings, hiPSC-CMs were dissociated with TrypLE Select 10X (Gibco) and plated on Matrigel (Corning) coated glass coverslips as single cells with a density of 8.4 - 10.5 * 10^3^/cm^2^.

**Action potential recording**

APs were recorded in perforated patch mode 3-10 days after hiPSC-CMs dissociation. The extracellular solution was based on modified Tyrode’s solution and contained (mM): NaCl 154, KCl 5.4, CaCl2 1.8, HEPES-NaOH 5, D-Glucose 5.5. pH was set to 7.35 with NaOH. The intracellular solution contained (mM): K-Aspartate 125, KCl 20, NaCl 10, Na2-ATP 5, HEPES 10. pH was set to 7.3 with KOH. Amphotericin B 0.22 mM, dissolved in DMSO, was added to the intracellular solution to record APs.

APs were recorded with a Molecular Devices digidata 1440A and a Molecular Devices Axopatch 200B amplifier at physiological temperature (37 °C) and under 1 Hz pacing. Current pulses with duration of 2-3 ms and amplitudes of 0.5-1.5 nA were used to elicit APs. APs were digitized at 5 kHz and filtered at 2 kHz with a low-pass Bessel filter. Liquid junction potential was calculated according to the stationary Nernst-Planck equation using LJPcalc (Harden, SW and Brogioli, D (2020). LJPcalc [Online]. Available: <https://swharden.com/software/LJPcalc>, Accessed on 03/08/2021). The calculated LJP was 13.922 mV. The measured LJP was -11.2 ± 0.9 mV.

#### **Slow delayed rectifier potassium current recording**

The slow delayed rectifier potassium current (I_Ks_) was recorded with the patch clamp technique in isolated hiPSC-CMs. Signals were digitized at 2 kHz and filtered at 1 kHz through a Molecular Device 1440A Digidata connected to a Molecular Devices 200B amplifier. Currents were recorded at physiological temperature (37 °C). The extracellular solution contained (mM): NaCl 154, KCl 5.4, CaCl2 1.8, HEPES-NaOH 5, D-Glucose 5.5; pH was set to 7.35 with NaOH. The intracellular solutions for I_Ks_ recordings contained (mM): K-Aspartate 110, KCl 23, MgCl2 3, EGTA-KOH 5, HEPES-KOH 5, Guanosine-5'-triphosphate disodium salt 0.4, Adenosine-5'-triphosphate disodium salt 5, Phosphatidylcholine sodium salt 5, and CaCl2 2. Currents were evoked from a holding potential of -40 mV, with depolarizing voltage steps of 4000 ms every 10 mV up to +60 mV, followed by 8000 ms at -40 mV. I_Ks_ was isolated as 1 µM HMR-1556 (Selleckchem) sensitive current, in the presence of 1 µM of the I_Kr_ blocker E4031 dihydrochloride (Selleckchem) and 1 µM of the L-type calcium channel blocker nifedipine (Merck). pClamp 10.4 was used to record the traces and Clampfit 10 was used for the analyses.

#### **MultiElectrode Arrays**

#### Multiwell MEAs (24 wells, MultiChannel Systems) were coated with 40 µg/mL bovine fibronectin (Merck) for 1h at 37 °C and processed as previously described ^4,5^. Expanded hiPSC-CMs were dissociated with TrypLE Select Enzyme 10X (Gibco) and plated on the electrodes of Multiwell MEAs at a density of 5 * 10^4^ hiPSC-CMs per well in small drops. RBK medium supplemented with Revitacell (Gibco) was used for plating. The next day, and thereafter twice a week, half of the medium was refreshed with RBK medium. After two weeks, baseline recordings were performed followed by acute drug treatment and cumulative concentration-response measurements.

##### *Drug testing*

The drug testing protocol included the following steps: 1) recording of baseline Field Potentials (FP), 2) addition of vehicle (RBK) medium to mimic temperature and mechanical impact of drug addiction 3) addition of cumulative doses of compounds . All the recordings simulated acute drug exposure and consisted in 1 minute of wash-in followed by two minutes of recording. The following drugs were tested: chlorpromazine hydrochloride (Selleckchem), ciprofloxacin hydrochloride hydrate (Selleckchem), clarithromycin (Selleckchem), dofetilide (Selleckchem), E4031 dihydrochloride (Selleckchem), haloperidol (Selleckchem), HMR-1556 (Selleckchem), moxifloxacin (Selleckchem), nifedipine (Merck), salbutamol sulfate (Selleckchem), tetrodotoxin citrate (Tocris).

##### *Field Potential analysis*

MEA recordings were analyzed using Multiwell-Analyzer v2.0.6.0 (Multichannel Systems) and the following parameters were extracted: field potential duration (FPD), beat-to-beat interval (RR), peak-to-peak amplitude (PtPA), mean slope of depolarization (Mean slope), RR interval coefficient of variation (RRCV). The analysis of FPs was performed as previously described ^4^ and automated through custom R scripts (R v4.3.1). FPD was corrected for beating frequency (cFPD) using Bazett’s formula.

##### *Field Potential quality assessment*

The quality of the FP was assessed with a scoring system as previously described ^5^ and then further stratified to four categories based on the beating patterns: normal beating (scores 3-5), irregular beating (score 2), arrhythmic beating (score 1), and quiescence (score 0). This was implemented with the rationale of separating drug effects on the quantitative properties of the FP (e.g. a FP prolongation) from the potential detrimental effects of the drug treatment on FP quality as no FPD was measured from recordings scored as 1 and 0.

### **Supplemental results**

#### **Generation of hiPSC lines**

Lines SA14.19 and SA15.14, carrying the *KCNQ1* A341V genetic variant, were generated from dermal fibroblasts via retroviral infection with *OCT4*, *SOX2*, *KLF4*, and *cMYC* and fully characterized (Figure S1) following the procedure described previously ^6,7^. Line 26.1, carrying the *KCNH2* R366X genetic variant, was generated from dermal fibroblasts through transfection with two oriP/EBNA1-based episomal plasmids and fully characterized following the previously described procedure ^8^.

### **Supplemental tables**

**Supplemental Table 1. List of hiPSC lines used in the study and their characterization status.**

|  | **Genetic variant** | **Known modifier gene** | **Disease** | **Line name** | **Line ID** | **Reference** |
| --- | --- | --- | --- | --- | --- | --- |
| 1 | Wild Type |  | Healthy | WTC-11 |  | ^9^ |
| 2 | *KCNQ1* p.R190W |  | LQT1 | 29.7 | PSMi005-A | ^10^ |
| 3 | *KCNQ1* p.R594Q |  | LQT1 | 28.44 | PSMi004-A | ^11^ |
| 4 | *KCNQ1* p.A341V |  | LQT1 | SA6.27 | PSMi001-A | ^12^ |
| 5 | *KCNQ1*  p.A341V |  | LQT1 | SA15.14 |  | Supplemental Figure 1 |
| 6 | *KCNQ*1 p.A341V | *NOS1AP* rs16847548 and rs4657139 heterozygous minor alleles | LQT1 | SA14.19 |  | Supplemental Figure 1 |
| 7 | *KCNQ1* p.A341V | *NOS1AP* rs16847548 and rs4657139 homozygous minor alleles | LQT1 | SA13.5 | PSMi007-A | ^7,12^ |
| 8 | *KCNQ1* p.R190W & *KCNQ1* p.R594Q |  | JLNS | 30.3 | PSMi002-A | ^13^ |
| 9 | *KCNH2* p.R366X |  | LQT2 | 26.1 |  | Supplemental Figure 2 |
| 10 | *KCNH2* p.R366X |  | LQT2 | 27.2 |  | ^8^ |
| 11 | *KCNH2* p.A561V |  | LQT2 | SCVI498c1 |  | ^14^ |

**Supplemental Table 2. List of main reagents used in the study.**

| **Reagent** | **Company** | **Product code** |
| --- | --- | --- |
| B-27™ Supplement | Thermo Fisher Scientific [GIBCO] | 17504044 |
| B-27™ Supplement, minus insulin | Thermo Fisher Scientific [GIBCO] | A1895601 |
| Bambanker | Nippon Genetics | BB03 |
| CHIR-99021 HCl | Selleckchem | S2924 |
| Chlorpromazine | SelleckChem | #S5749 |
| Ciprofloxacin hydrochloride hydrate | SelleckChem | #S5208 |
| Clarithromycin | SelleckChem | #S2555 |
| Dofetilide | SelleckChem | #S1658 |
| DPBS, no calcium, no magnesium | Thermo Fisher Scientific [GIBCO] | 14190094 |
| E4031 dihydrochloride | Tocris | #1808 |
| EDTA (0.5 M), pH 8.0, RNase-free | Thermo Fisher Scientific [Invitrogen] | AM9262 |
| Essential 8™ Flex Medium Kit | Thermo Fisher Scientific [GIBCO] | A2858501 |
| Fibronectin bovine plasma | Merck [Sigma-Aldrich] | F1141 |
| Haloperidol | SelleckChem | #S1920 |
| HMR1556 | Tocris | #5011 |
| IWR-1 | Merck [Sigma-Aldrich] | I0161 |
| KOSR | Thermo Fisher Scientific 10828028  [GIBCO] | |
| Lactic acid | Merck | 27714 |
| Matrigel® Corning® hESC-Qualified Matrix, LDEV-free | Corning | 354277 |
| Moxifloxacin | SelleckChem | #S5535 |
| Nifedipine | Merck | #N7634 |
| RevitaCell™ Supplement | Thermo Fisher Scientific [GIBCO] | A2644501 |
| RPMI 1640 Medium, no glucose | Thermo Fisher Scientific [GIBCO] | 11879020 |
| RPMI 1640 w/ L-Glutamine | Euroclone | ECB2000 |
| Salbutamol | SelleckChem | #S2507 |
| Tetrodotoxin citrate | Tocris | #1069 |
| TrypLE™ Select Enzyme | Thermo Fisher Scientific [GIBCO] | A1217701 |
| Vitronectin (VTN-N) Recombinant Human Protein, Truncated | Thermo Fisher Scientific [GIBCO] | A14700 |

**Supplemental Table 3. List of pro-arrhythmic compounds in the CredibleMeds® database associated with specific OpenFDA queries.**

| **Drug** | **Drug Class** | **Therapeutic Use** | **Classification on CredibleMeds** |
| --- | --- | --- | --- |
| albuterol (salbutamol) | Bronchodilator | Asthma | Avoid in congenital long QT |
| amiodarone | Antiarrhythmic | Arrhythmia | Risk of TdPAnd Avoid in congenital long QT |
| amitriptyline | Antidepressant, Tricyclic | Depression | Conditional Risk of TdPAnd Avoid in congenital long QT |
| amphotericin b | Antifungal | Fungal infection | Conditional Risk of TdPAnd Avoid in congenital long QT |
| aripiprazole | Antipsychotic, atypical | Schizophrenia, depression (adjunct) | Possible Risk of TdPAnd Avoid in congenital long QT |
| chlorpromazine | Antipsychotic, antiemetic | Schizophrenia, bipolar disorder, acute psychosis | Risk of TdPAnd Avoid in congenital long QT |
| cimetidine | H2-receptor antagonist | Gastric hyperacidity, GERD | Conditional Risk of TdPAnd Avoid in congenital long QT |
| ciprofloxacin | Antibiotic | Bacterial infection | Risk of TdPAnd Avoid in congenital long QT |
| clarithromycin | Antibiotic | Bacterial infection | Risk of TdPAnd Avoid in congenital long QT |
| clofazimine | Antibiotic | Leprosy | Possible Risk of TdPAnd Avoid in congenital long QT |
| clozapine | Antipsychotic, atypical | Schizophrenia | Possible Risk of TdPAnd Avoid in congenital long QT |
| diphenhydramine | Antihistamine | Allergic rhinitis, insomnia | Conditional Risk of TdPAnd Avoid in congenital long QT |
| dofetilide | Antiarrhythmic | Arrhythmia | Risk of TdPAnd Avoid in congenital long QT |
| donepezil | Cholinesterase inhibitor | Dementia (Alzheimer's Disease) | Risk of TdPAnd Avoid in congenital long QT |
| famotidine | H2-receptor antagonist | Gastric hyperacidity, GERD | Conditional Risk of TdPAnd Avoid in congenital long QT |
| flecainide | Antiarrhythmic | Arrhythmia | Risk of TdPAnd Avoid in congenital long QT |
| fluconazole | Antifungal | Fungal infection | Risk of TdPAnd Avoid in congenital long QT |
| fluorouracil | Anti-cancer | Cancer | Possible Risk of TdPAnd Avoid in congenital long QT |
| furosemide | Diuretic | Hypertension, diuresis | Conditional Risk of TdPAnd Avoid in congenital long QT |
| haloperidol | Antipsychotic | Schizophrenia, agitation | Risk of TdPAnd Avoid in congenital long QT |
| hydrochlorothiazide | Diuretic | Hypertension, diuresis | Conditional Risk of TdPAnd Avoid in congenital long QT |
| hydroxychloroquine | Antimalarial, Anti-inflammatory | Malaria, SLE, rheumatoid arthritis | Conditional Risk of TdPAnd Avoid in congenital long QT |
| indapamide | Diuretic | Hypertension, diuresis | Conditional Risk of TdPAnd Avoid in congenital long QT |
| ivabradine | Antianginal | Angina Pectoris (heart pain) | Conditional Risk of TdPAnd Avoid in congenital long QT |
| lansoprazole | Proton Pump Inhibitor | Gastric hyperacidity, GERD | Conditional Risk of TdPAnd Avoid in congenital long QT |
| levofloxacin | Antibiotic | Bacterial infection | Risk of TdPAnd Avoid in congenital long QT |
| loperamide | Opiate | Diarrhea | Conditional Risk of TdPAnd Avoid in congenital long QT |
| metoclopramide | Antiemetic | Nausea, vomiting | Conditional Risk of TdPAnd Avoid in congenital long QT |
| metronidazole | Antibiotic | Trichomoniasis, amebiasis, bacterial infection | Conditional Risk of TdPAnd Avoid in congenital long QT |
| mirtazapine | Antidepressant, Tetracyclic | Depression | Possible Risk of TdPAnd Avoid in congenital long QT |
| moxifloxacin | Antibiotic | Bacterial infection | Risk of TdPAnd Avoid in congenital long QT |
| nilotinib | Anti-cancer | Cancer (leukemia) | Possible Risk of TdPAnd Avoid in congenital long QT |
| olanzapine | Antipsychotic, atypical | Schizophrenia, bipolar disorder | Conditional Risk of TdPAnd Avoid in congenital long QT |
| omeprazole | Proton Pump Inhibitor | Gastric hyperacidity, GERD | Conditional Risk of TdPAnd Avoid in congenital long QT |
| ondansetron | Antiemetic | Nausea, vomiting | Risk of TdPAnd Avoid in congenital long QT |
| pantoprazole | Proton Pump Inhibitor | Gastric hyperacidity, GERD | Conditional Risk of TdPAnd Avoid in congenital long QT |
| propofol | Anesthetic, general | Anesthesia | Risk of TdPAnd Avoid in congenital long QT |
| quetiapine | Antipsychotic, atypical | Schizophrenia | Conditional Risk of TdPAnd Avoid in congenital long QT |
| risperidone | Antipsychotic, atypical | Schizophrenia | Possible Risk of TdPAnd Avoid in congenital long QT |
| sotalol | Antiarrhythmic | Arrhythmia | Risk of TdPAnd Avoid in congenital long QT |
| sulpiride | Antipsychotic, atypical | Schizophrenia | Risk of TdPAnd Avoid in congenital long QT |
| tacrolimus | Immunosuppressant | Immune suppression | Possible Risk of TdPAnd Avoid in congenital long QT |
| torsemide | Diuretic | Hypertension, diuresis | Possible Risk of TdPAnd Avoid in congenital long QT |
| tramadol | Analgesic | Pain | Possible Risk of TdPAnd Avoid in congenital long QT |
| voriconazole | Antifungal | Fungal infection | Conditional Risk of TdPAnd Avoid in congenital long QT |

Compounds selected for the study are highlighted in pink.

**Supplemental Table 4. Ion channel blockers and pro-arrhythmic compounds used in the study.**

| **Compound** | **Drug class** | **Therapeutic use** | **IC50 HERG** | **Cmax** | **Tested concentration range** | **Stock concentration** | **Vehicle** |
| --- | --- | --- | --- | --- | --- | --- | --- |
| Chlorpromazine | Antipsychotic, antiemetic | Schizophrenia, bipolar disorder, acute psychosis | 21.6 μM ^15^ | 0.0345 µM ^16^ | 0.03 - 3 µM | 100 mM | DMSO |
| Ciprofloxacin | Antibiotic | Bacterial  infection | 966 μM ^17^ | 8.87 µM ^18^ | 3 - 30 µM | 15 mM | DMSO |
| Clarithromycin | Antibiotic | Bacterial infection | 45.7 μM ^19^ | 1.206 µM ^16^ | 1 - 10 µM | 6 mM | DMSO |
| Dofetilide | Antiarrhythmic | Arrhythmia | 0.007 μM ^20^ | 0.0023 µM ^16^ | 1 - 100 nM | 50 mM | DMSO |
| E4031 | Selective I_Kr_ blocker | - | 0.0077 μM ^21^ | - | 10 - 500 nM | 10 mM | Water |
| Haloperidol | Antipsychotic | Schizophrenia, agitation | 1 μm ^22^ | 0.079 µM ^23^ | 0.3 - 10 µM | 150 mM | DMSO |
| HMR1556 | Selective I_Ks_ blocker | - | - | - | 0.3 - 10 µM | 10 mM | DMSO |
| Moxifloxacin | Antibiotic | Bacterial infection | 41.2 μM ^24^ | 11.7 μM ^25^ | 1 - 100 µM | 100 mM | DMSO |
| Nifedipine | Selective I_CaL_ blocker | Angina, high blood pressure | - | 0.0077 µM ^16^ | 0.03 - 2 µM | 10 mM | Ethanol, 100% |
| Salbutamol (albuterol) | Bronchodilator | Asthma | - | 0.059 µM ^26^ | 0.003 - 0.3 µM | 100 mM | Water |
| Tetrodotoxin | Selective I_Na_ blocker | - | - | - | 0.1 - 30 µM | 3.13 mM | Water |

**Supplemental Table 5. Mean patients’ QTc values and mean Schwartz scores for each genetic variant included in the study.**

| **Variant** | **N** | **QTc (ms)** | **Schwartz Score** |
| --- | --- | --- | --- |
| KCNQ1-p.R190W | 102 | 464.7 ± 38 | 2.9 ± 1.5 |
| KCNQ1-p.R594Q | 16 | 491.9 ± 61.3 | 3.2 ± 2.2 |
| KCNQ1-p.R190W & p.R594Q | 1 | 578 | 7.5 |
| KCNQ1-p.A341V | 183 | 489.9 ± 45.8 | 4.7 ± 1.6 |
| KCNH2-p.R366X | 12 | 501.2 ± 56.9 | 4.5 ± 1.9 |
| KCNH2-p.A561V | 13 | 543.4 ± 34.6 | 5.0 ± 1.0 |

Data is presented as mean ± standard deviation. N indicates the number of patients included in the analysis.

**Supplemental Table 6. Genetic variants selected for the study and their functional characterisation status.**

| **Genetic variant** | **Disease** | **dbSNP record** | **ClinVar classification** | **Functional characterization** | **Reference** |
| --- | --- | --- | --- | --- | --- |
| *KCNQ1* p.R190W | LQT1 | [rs199473662](https://www.ncbi.nlm.nih.gov/snp/rs199473662) | Likely pathogenic | This paper |  |
| *KCNQ1* p.R594Q | LQT1 | [rs199472815](https://www.ncbi.nlm.nih.gov/snp/rs199472815) | Likely pathogenic | loss of function, trafficking defect | ^27^ |
| *KCNQ1* p.A341V | LQT1 | [rs12720459](https://www.ncbi.nlm.nih.gov/snp/rs12720459) | Pathogenic | loss of function | ^28–30^ |
| *KCNQ1* p.R190W & *KCNQ1* p.R594Q | JLNS | [rs199473662](https://www.ncbi.nlm.nih.gov/snp/rs199473662),  [rs199472815](https://www.ncbi.nlm.nih.gov/snp/rs199472815) | - | This paper |  |
| *KCNH2* p.R366X | LQT2 | [rs794728364](https://www.ncbi.nlm.nih.gov/snp/rs794728364) | Pathogenic | FPD prolongation (iPSC-CMs) | ^8^ |
| *KCNH2* p.A561V | LQT2 | [rs121912504](https://www.ncbi.nlm.nih.gov/snp/rs121912504) | Pathogenic/Likely pathogenic | loss of function, trafficking defect | ^14,31–34^ |

**Supplemental Table 7. Differential response of Low Risk and High Risk hiPSC-CMs to ion channel blockers and proarrhythmic drugs.**

| **Variant Risk Level** | **Drug** | **Dose** | **N** | **Normalized FPD (% of Vehicle)** | **Normalized cFPD (% of Vehicle)** | **Normalized RR (% of Vehicle)** | **Normalized PtPA (% of Vehicle)** |
| --- | --- | --- | --- | --- | --- | --- | --- |
| Low Risk | Chlorpromazine | 30 nM | 32 | 108.7 ± 7, p = 0.01 | 113.7 ± 16.2, p = 0.58 | 93.7 ± 13.8, p = 0.94 | 99.6 ± 15.5, p = 0.27 |
| Low Risk | Chlorpromazine | 100 nM | 32 | 113.2 ± 10.5, p = 0.01 | 122.9 ± 24.5, p = 0.78 | 88.6 ± 15.8, p = 0.26 | 98.2 ± 20.6, p = 0.17 |
| Low Risk | Chlorpromazine | 300 nM | 32 | 116.4 ± 12.2, p ≤ 0.001 | 129.6 ± 27.4, p = 0.06 | 84.5 ± 15.2, p = 0.1 | 97.8 ± 23.9, p = 0.68 |
| Low Risk | Chlorpromazine | 1 µM | 32 | 118.5 ± 13.2, p ≤ 0.001 | 134.6 ± 28.5, p = 0.005 | 80.9 ± 13.7, p = 0.01 | 92.7 ± 37.2, p = 0.52 |
| Low Risk | Chlorpromazine | 3 µM | 32 | 117 ± 18.6, p ≤ 0.001 | 138.1 ± 35.7, p ≤ 0.001 | 75.8 ± 14.7, p = 0.08 | 67.5 ± 34.5, p = 0.15 |
| High Risk | Chlorpromazine | 30 nM | 67 | 105.8 ± 4.3, p = 0.01 | 108.9 ± 7.8, p = 0.58 | 95.4 ± 12, p = 0.94 | 101.5 ± 16.1, p = 0.27 |
| High Risk | Chlorpromazine | 100 nM | 67 | 106.8 ± 6.3, p = 0.01 | 114.9 ± 9.9, p = 0.78 | 87.9 ± 13.3, p = 0.26 | 101.7 ± 20.7, p = 0.17 |
| High Risk | Chlorpromazine | 300 nM | 67 | 105.6 ± 8.6, p ≤ 0.001 | 117.7 ± 12.6, p = 0.06 | 82.7 ± 18.6, p = 0.1 | 97.1 ± 24, p = 0.68 |
| High Risk | Chlorpromazine | 1 µM | 67 | 101 ± 12.6, p ≤ 0.001 | 117.7 ± 16.3, p = 0.005 | 76 ± 21.6, p = 0.01 | 90.9 ± 24.2, p = 0.52 |
| High Risk | Chlorpromazine | 3 µM | 67 | 93.7 ± 17.6, p ≤ 0.001 | 112.9 ± 23, p ≤ 0.001 | 72.2 ± 27, p = 0.08 | 74.2 ± 29.1, p = 0.15 |
| Low Risk | Ciprofloxacin | 3 µM | 56 | 106.9 ± 3.3, p = 0.59 | 106.9 ± 5.6, p = 0.01 | 100.5 ± 8.1, p = 0.003 | 103.4 ± 21.7, p = 0.14 |
| Low Risk | Ciprofloxacin | 10 µM | 56 | 110 ± 6.1, p = 0.28 | 111.2 ± 8, p = 0.54 | 98.6 ± 10.1, p = 0.14 | 101.5 ± 20.5, p = 0.28 |
| Low Risk | Ciprofloxacin | 30 µM | 56 | 95.3 ± 9.3, p ≤ 0.001 | 104.7 ± 9.4, p = 0.92 | 83.2 ± 8.9, p ≤ 0.001 | 106.8 ± 46.3, p = 0.68 |
| High Risk | Ciprofloxacin | 3 µM | 104 | 106.6 ± 5.5, p = 0.59 | 110.1 ± 9.5, p = 0.01 | 95.3 ± 14.2, p = 0.003 | 98.2 ± 16.3, p = 0.14 |
| High Risk | Ciprofloxacin | 10 µM | 104 | 108.8 ± 7.9, p = 0.28 | 113.5 ± 13.3, p = 0.54 | 94.3 ± 16.7, p = 0.14 | 98.1 ± 22.9, p = 0.28 |
| High Risk | Ciprofloxacin | 30 µM | 104 | 90.2 ± 10.5, p ≤ 0.001 | 107.1 ± 16.7, p = 0.92 | 73.2 ± 14.5, p ≤ 0.001 | 103.3 ± 27.7, p = 0.68 |
| Low Risk | Clarithromycin | 1 µM | 57 | 108.6 ± 4.7, p = 0.09 | 109.8 ± 6, p = 0.79 | 98.3 ± 8.7, p = 0.27 | 100.3 ± 14.3, p = 0.17 |
| Low Risk | Clarithromycin | 3 µM | 57 | 113 ± 10, p = 0.09 | 120.1 ± 9.6, p = 0.99 | 89 ± 10.5, p = 0.27 | 96.8 ± 16.3, p = 0.02 |
| Low Risk | Clarithromycin | 10 µM | 57 | 64.6 ± 27.4, p = 0.54 | 89.5 ± 28.2, p = 0.12 | 55.1 ± 22.8, p = 0.03 | 43.5 ± 44.8, p = 0.74 |
| High Risk | Clarithromycin | 1 µM | 111 | 106.6 ± 6.3, p = 0.09 | 109.4 ± 7.3, p = 0.79 | 95.9 ± 12.8, p = 0.27 | 96.8 ± 9.3, p = 0.17 |
| High Risk | Clarithromycin | 3 µM | 111 | 110.3 ± 11.4, p = 0.09 | 120.6 ± 15.8, p = 0.99 | 86.3 ± 19.4, p = 0.27 | 90.3 ± 14.6, p = 0.02 |
| High Risk | Clarithromycin | 10 µM | 111 | 74.8 ± 79.5, p = 0.54 | 93.2 ± 78.8, p = 0.12 | 88.3 ± 136.7, p = 0.03 | 41.8 ± 36.1, p = 0.74 |
| Low Risk | Dofetilide | 1 nM | 64 | 109.7 ± 3.2, p = 0.22 | 108.4 ± 4.1, p = 0.002 | 102.8 ± 7.1, p = 0.02 | 98.4 ± 12.1, p = 0.49 |
| Low Risk | Dofetilide | 3 nM | 64 | 125.2 ± 8.3, p = 0.002 | 124.4 ± 8.8, p ≤ 0.001 | 101.8 ± 9.4, p = 0.04 | 95.9 ± 18, p = 0.97 |
| Low Risk | Dofetilide | 10 nM | 64 | 158.9 ± 23.8, p ≤ 0.001 | 153.8 ± 20.8, p ≤ 0.001 | 107.5 ± 17.3, p = 0.73 | 88.4 ± 23.8, p = 0.09 |
| Low Risk | Dofetilide | 30 nM | 64 | 201.3 ± 54.8, p ≤ 0.001 | 188.4 ± 38, p ≤ 0.001 | 114.6 ± 26.2, p = 0.24 | 81.5 ± 23, p = 0.005 |
| Low Risk | Dofetilide | 100 nM | 64 | 224.3 ± 94.8, p ≤ 0.001 | 208.2 ± 61.4, p ≤ 0.001 | 134.9 ± 71.9, p = 0.4 | 65 ± 27.4, p = 0.03 |
| High Risk | Dofetilide | 1 nM | 121 | 111.1 ± 7, p = 0.22 | 111.9 ± 9.9, p = 0.002 | 100.2 ± 16.3, p = 0.02 | 99.7 ± 8.8, p = 0.49 |
| High Risk | Dofetilide | 3 nM | 121 | 133.2 ± 19.3, p = 0.002 | 135.4 ± 17.5, p ≤ 0.001 | 98.1 ± 18.3, p = 0.04 | 95.5 ± 15.6, p = 0.97 |
| High Risk | Dofetilide | 10 nM | 121 | 202.4 ± 55.8, p ≤ 0.001 | 192.1 ± 45.3, p ≤ 0.001 | 128.2 ± 193.3, p = 0.73 | 84.3 ± 21.5, p = 0.09 |
| High Risk | Dofetilide | 30 nM | 121 | 298.9 ± 114.8, p ≤ 0.001 | 269.6 ± 101.7, p ≤ 0.001 | 132.4 ± 80.5, p = 0.24 | 68.5 ± 30.6, p = 0.005 |
| High Risk | Dofetilide | 100 nM | 121 | 374.1 ± 153.7, p ≤ 0.001 | 317.5 ± 128.5, p ≤ 0.001 | 156.4 ± 142.2, p = 0.4 | 54.6 ± 32.4, p = 0.03 |
| Low Risk | E4031 | 10 nM | 48 | 121.8 ± 16.6, p = 0.75 | 122.1 ± 14.8, p = 0.96 | 99.8 ± 10.4, p = 0.6 | 100.1 ± 24.1, p = 0.21 |
| Low Risk | E4031 | 30 nM | 48 | 156.7 ± 27.3, p ≤ 0.001 | 157.3 ± 26.8, p = 0.003 | 100.3 ± 15.2, p = 0.009 | 93.9 ± 32.7, p = 0.27 |
| Low Risk | E4031 | 100 nM | 48 | 192.8 ± 49.5, p ≤ 0.001 | 191.3 ± 47.9, p ≤ 0.001 | 102.3 ± 15.9, p = 0.01 | 85 ± 41.7, p = 0.07 |
| Low Risk | E4031 | 300 nM | 48 | 219 ± 66.2, p ≤ 0.001 | 213.7 ± 58.9, p ≤ 0.001 | 105.6 ± 21.8, p ≤ 0.001 | 70.8 ± 49.1, p = 0.25 |
| Low Risk | E4031 | 500 nM | 48 | 226.2 ± 70.7, p ≤ 0.001 | 218.2 ± 69.9, p ≤ 0.001 | 126 ± 143.4, p = 0.003 | 64.4 ± 51.7, p = 0.08 |
| High Risk | E4031 | 10 nM | 77 | 128.3 ± 35, p = 0.75 | 126.8 ± 28.4, p = 0.96 | 101.8 ± 13.8, p = 0.6 | 97 ± 12.6, p = 0.21 |
| High Risk | E4031 | 30 nM | 77 | 204 ± 80.4, p ≤ 0.001 | 192.5 ± 78.7, p = 0.003 | 114.2 ± 28.1, p = 0.009 | 87.5 ± 16.3, p = 0.27 |
| High Risk | E4031 | 100 nM | 77 | 290.4 ± 186.5, p ≤ 0.001 | 253.8 ± 129.3, p ≤ 0.001 | 124.4 ± 46.6, p = 0.01 | 71.1 ± 27.4, p = 0.07 |
| High Risk | E4031 | 300 nM | 77 | 374.1 ± 215.6, p ≤ 0.001 | 314 ± 166.4, p ≤ 0.001 | 148 ± 73, p ≤ 0.001 | 59.4 ± 25, p = 0.25 |
| High Risk | E4031 | 500 nM | 73 | 413.6 ± 245.1, p ≤ 0.001 | 350 ± 183.2, p ≤ 0.001 | 162.6 ± 166.1, p = 0.003 | 48.3 ± 29.8, p = 0.08 |
| Low Risk | Haloperidol | 300 nM | 57 | 139.4 ± 32.3, p ≤ 0.001 | 129.8 ± 27.1, p ≤ 0.001 | 115.6 ± 18.9, p = 0.06 | 93.4 ± 21.9, p = 0.21 |
| Low Risk | Haloperidol | 1 µM | 57 | 204.7 ± 76.2, p ≤ 0.001 | 178.2 ± 50.6, p ≤ 0.001 | 132.4 ± 43.4, p = 0.007 | 69 ± 30, p = 0.72 |
| Low Risk | Haloperidol | 3 µM | 57 | 200.1 ± 64, p ≤ 0.001 | 177.8 ± 40.1, p ≤ 0.001 | 126.7 ± 42.7, p = 0.05 | 49.5 ± 31.7, p = 0.86 |
| Low Risk | Haloperidol | 10 µM | 57 | 163.8 ± 48.1, p = 0.003 | 150.5 ± 31.8, p ≤ 0.001 | 124.7 ± 62.1, p = 0.63 | 41.1 ± 29.8, p = 0.06 |
| High Risk | Haloperidol | 300 nM | 118 | 203.4 ± 143.1, p ≤ 0.001 | 180.1 ± 109, p ≤ 0.001 | 123.5 ± 33.3, p = 0.06 | 91.8 ± 16.1, p = 0.21 |
| High Risk | Haloperidol | 1 µM | 118 | 306.9 ± 180.3, p ≤ 0.001 | 253.1 ± 134, p ≤ 0.001 | 149.2 ± 65.7, p = 0.007 | 71.9 ± 26.1, p = 0.72 |
| High Risk | Haloperidol | 3 µM | 118 | 287.2 ± 148.6, p ≤ 0.001 | 251 ± 125.2, p ≤ 0.001 | 139.8 ± 59.9, p = 0.05 | 47.4 ± 28.1, p = 0.86 |
| High Risk | Haloperidol | 10 µM | 118 | 236.9 ± 169.1, p = 0.003 | 212.9 ± 126, p ≤ 0.001 | 144.6 ± 128.6, p = 0.63 | 32.4 ± 26.3, p = 0.06 |
| Low Risk | HMR1556 | 300 nM | 43 | 111.4 ± 5.8, p = 0.006 | 115 ± 14.1, p = 0.43 | 95.4 ± 10.8, p = 0.002 | 100.1 ± 24.5, p = 0.21 |
| Low Risk | HMR1556 | 1 µM | 43 | 112.9 ± 7.3, p ≤ 0.001 | 120.2 ± 15.5, p = 0.65 | 89.8 ± 10.7, p = 0.005 | 96.8 ± 26.9, p = 0.15 |
| Low Risk | HMR1556 | 3 µM | 43 | 112.9 ± 9.5, p ≤ 0.001 | 123.3 ± 18.6, p = 0.05 | 85.6 ± 12, p ≤ 0.001 | 92.3 ± 25.2, p = 0.17 |
| Low Risk | HMR1556 | 10 µM | 43 | 89.4 ± 19.7, p ≤ 0.001 | 107.3 ± 27.3, p = 0.04 | 70.8 ± 12.9, p ≤ 0.001 | 80.8 ± 43.9, p = 0.87 |
| High Risk | HMR1556 | 300 nM | 75 | 107.8 ± 7.6, p = 0.006 | 116.9 ± 17.5, p = 0.43 | 88.5 ± 18.6, p = 0.002 | 96.9 ± 11.8, p = 0.21 |
| High Risk | HMR1556 | 1 µM | 75 | 106.2 ± 8.1, p ≤ 0.001 | 120.1 ± 21.5, p = 0.65 | 82.1 ± 17.7, p = 0.005 | 92.7 ± 15.7, p = 0.15 |
| High Risk | HMR1556 | 3 µM | 75 | 99.4 ± 9.6, p ≤ 0.001 | 120 ± 25.1, p = 0.05 | 73.2 ± 18.9, p ≤ 0.001 | 88.1 ± 16.2, p = 0.17 |
| High Risk | HMR1556 | 10 µM | 72 | 74 ± 18.7, p ≤ 0.001 | 97.5 ± 26.2, p = 0.04 | 63.8 ± 46.5, p ≤ 0.001 | 75.9 ± 21.5, p = 0.87 |
| Low Risk | HMR1556 (after E4031) | E4031  20 nM | 46 | 135.3 ± 21.7, p = 0.84 | 131 ± 21.3, p = 0.83 | 108.3 ± 18.6, p = 0.91 | 94.1 ± 10.8, p = 0.07 |
| Low Risk | HMR1556 (after E4031) | 300 nM | 46 | 163.1 ± 25.9, p ≤ 0.001 | 159 ± 23.2, p = 0.001 | 106.1 ± 15.6, p = 0.08 | 85.8 ± 18, p = 0.28 |
| Low Risk | HMR1556 (after E4031) | 1 µM | 46 | 167.7 ± 29.2, p = 0.003 | 167.4 ± 26.5, p = 0.004 | 101.1 ± 15.1, p = 0.9 | 80 ± 21.1, p = 0.32 |
| Low Risk | HMR1556 (after E4031) | 3 µM | 46 | 163.7 ± 26.3, p = 0.37 | 169.2 ± 26.4, p = 0.25 | 94.4 ± 12.8, p = 0.08 | 75.7 ± 21.3, p = 0.39 |
| Low Risk | HMR1556 (after E4031) | 10 µM | 46 | 107.1 ± 21.4, p = 0.2 | 128.6 ± 26.6, p = 0.8 | 70.3 ± 11.2, p = 0.06 | 71.9 ± 34.3, p = 0.39 |
| High Risk | HMR1556 (after E4031) | E4031  20 nM | 73 | 151.1 ± 61.1, p = 0.84 | 140.2 ± 41, p = 0.83 | 114.5 ± 31.2, p = 0.91 | 96.7 ± 10.5, p = 0.07 |
| High Risk | HMR1556 (after E4031) | 300 nM | 73 | 196.5 ± 53.4, p ≤ 0.001 | 183.2 ± 42.6, p = 0.001 | 116.4 ± 30.1, p = 0.08 | 90.8 ± 17.6, p = 0.28 |
| High Risk | HMR1556 (after E4031) | 1 µM | 73 | 190.6 ± 43.8, p = 0.003 | 188.4 ± 40.6, p = 0.004 | 105.2 ± 28.7, p = 0.9 | 85.2 ± 18.4, p = 0.32 |
| High Risk | HMR1556 (after E4031) | 3 µM | 73 | 169.5 ± 34.4, p = 0.37 | 180.2 ± 42.7, p = 0.25 | 92.8 ± 26.7, p = 0.08 | 79.6 ± 20.8, p = 0.39 |
| High Risk | HMR1556 (after E4031) | 10 µM | 73 | 102.6 ± 29.4, p = 0.2 | 128.1 ± 35.9, p = 0.8 | 66.3 ± 18.4, p = 0.06 | 67.3 ± 22.1, p = 0.39 |
| Low Risk | Moxifloxacin | 1 µM | 44 | 106.2 ± 4.8, p = 0.07 | 108.2 ± 7.9, p = 0.78 | 97.6 ± 11.8, p = 0.53 | 96.6 ± 18.7, p = 0.5 |
| Low Risk | Moxifloxacin | 3 µM | 44 | 109.5 ± 8.3, p = 0.03 | 114.5 ± 15, p = 0.71 | 93.3 ± 12.8, p = 0.1 | 94.6 ± 16.5, p = 0.4 |
| Low Risk | Moxifloxacin | 10 µM | 44 | 114 ± 10.4, p = 0.007 | 122 ± 22.4, p = 0.98 | 90.1 ± 13.1, p = 0.03 | 92.8 ± 16, p = 0.61 |
| Low Risk | Moxifloxacin | 30 µM | 44 | 122.7 ± 12.5, p = 0.02 | 132 ± 27.2, p = 0.99 | 89.7 ± 13.7, p = 0.006 | 90.3 ± 14.8, p = 0.39 |
| Low Risk | Moxifloxacin | 100 µM | 43 | 260.7 ± 72.3, p = 0.005 | 236.6 ± 73.8, p = 0.04 | 137.9 ± 60.2, p ≤ 0.001 | 24.2 ± 36.1, p = 0.05 |
| High Risk | Moxifloxacin | 1 µM | 68 | 104.8 ± 4.9, p = 0.07 | 108.2 ± 9.2, p = 0.78 | 96 ± 18.6, p = 0.53 | 99 ± 8.9, p = 0.5 |
| High Risk | Moxifloxacin | 3 µM | 68 | 106.5 ± 7.8, p = 0.03 | 114.8 ± 13.8, p = 0.71 | 88.7 ± 16.6, p = 0.1 | 98.1 ± 9, p = 0.4 |
| High Risk | Moxifloxacin | 10 µM | 68 | 108.8 ± 11.1, p = 0.007 | 123.4 ± 26.5, p = 0.98 | 82.7 ± 19.3, p = 0.03 | 93.4 ± 10.4, p = 0.61 |
| High Risk | Moxifloxacin | 30 µM | 68 | 116.6 ± 18, p = 0.02 | 134.3 ± 32.8, p = 0.99 | 80.5 ± 20, p = 0.006 | 88.1 ± 21.5, p = 0.39 |
| High Risk | Moxifloxacin | 100 µM | 68 | 480.7 ± 277.4, p = 0.005 | 357.1 ± 199.6, p = 0.04 | 331.4 ± 452.4, p ≤ 0.001 | 21.8 ± 14.9, p = 0.05 |
| Low Risk | Nifedipine | 30 nM | 49 | 97.3 ± 7.6, p = 0.02 | 99.8 ± 8.6, p = 0.006 | 95.8 ± 13.6, p = 0.29 | 101.5 ± 16.1, p = 0.62 |
| Low Risk | Nifedipine | 100 nM | 49 | 81.7 ± 15.9, p = 0.07 | 89.6 ± 14.8, p = 0.17 | 86.7 ± 28.4, p = 0.008 | 97.8 ± 22.1, p = 1 |
| Low Risk | Nifedipine | 300 nM | 49 | 64.7 ± 19.1, p = 0.01 | 77.2 ± 18.4, p = 0.02 | 68.9 ± 11.4, p = 0.01 | 94.5 ± 23.2, p = 0.73 |
| Low Risk | Nifedipine | 1 µM | 49 | 50.5 ± 21, p = 0.003 | 64.8 ± 21.5, p = 0.007 | 58.3 ± 12.1, p ≤ 0.001 | 91.5 ± 23, p = 0.77 |
| Low Risk | Nifedipine | 2 µM | 49 | 35.6 ± 16.1, p ≤ 0.001 | 50.9 ± 18.4, p ≤ 0.001 | 47.1 ± 11.4, p ≤ 0.001 | 87.6 ± 29, p = 0.81 |
| High Risk | Nifedipine | 30 nM | 80 | 91.5 ± 17.1, p = 0.02 | 94.1 ± 13.8, p = 0.006 | 94.8 ± 23.7, p = 0.29 | 99.5 ± 16.9, p = 0.62 |
| High Risk | Nifedipine | 100 nM | 80 | 75.2 ± 17.3, p = 0.07 | 85.4 ± 17.6, p = 0.17 | 78.5 ± 22.5, p = 0.008 | 97 ± 20.7, p = 1 |
| High Risk | Nifedipine | 300 nM | 80 | 55.5 ± 19.4, p = 0.01 | 68.7 ± 20.5, p = 0.02 | 65.9 ± 30.9, p = 0.01 | 92.3 ± 22.6, p = 0.73 |
| High Risk | Nifedipine | 1 µM | 80 | 39.4 ± 17.9, p = 0.003 | 54.1 ± 19.1, p = 0.007 | 52 ± 22.9, p ≤ 0.001 | 89 ± 24.8, p = 0.77 |
| High Risk | Nifedipine | 2 µM | 76 | 26.2 ± 13.7, p ≤ 0.001 | 41.1 ± 19.2, p ≤ 0.001 | 39.2 ± 8, p ≤ 0.001 | 86 ± 24.9, p = 0.81 |
| Low Risk | Salbutamol | 3 nM | 41 | 104.3 ± 4.9, p ≤ 0.001 | 107.6 ± 7.8, p = 0.7 | 94.4 ± 7.7, p ≤ 0.001 | 98.8 ± 14.5, p = 1 |
| Low Risk | Salbutamol | 10 nM | 41 | 98.7 ± 9.6, p ≤ 0.001 | 108 ± 14.3, p = 0.73 | 84.6 ± 9, p ≤ 0.001 | 101.2 ± 16.7, p = 0.74 |
| Low Risk | Salbutamol | 30 nM | 41 | 86.5 ± 11.5, p ≤ 0.001 | 101.9 ± 16.5, p = 0.62 | 73.3 ± 10, p ≤ 0.001 | 102.4 ± 22.3, p = 0.2 |
| Low Risk | Salbutamol | 100 nM | 41 | 72.5 ± 10.4, p = 0.001 | 90.4 ± 14.1, p = 0.44 | 65.3 ± 9.4, p ≤ 0.001 | 102.7 ± 30.1, p = 0.33 |
| Low Risk | Salbutamol | 300 nM | 41 | 68.7 ± 7.9, p ≤ 0.001 | 87.1 ± 13.1, p = 0.97 | 67.2 ± 40.6, p ≤ 0.001 | 99.6 ± 26.3, p = 0.21 |
| High Risk | Salbutamol | 3 nM | 72 | 99.5 ± 8.9, p ≤ 0.001 | 109.2 ± 14.7, p = 0.7 | 85.4 ± 14.9, p ≤ 0.001 | 100.7 ± 14.3, p = 1 |
| High Risk | Salbutamol | 10 nM | 72 | 91.6 ± 11.2, p ≤ 0.001 | 110.9 ± 21.5, p = 0.73 | 71.9 ± 16.5, p ≤ 0.001 | 103.1 ± 17.4, p = 0.74 |
| High Risk | Salbutamol | 30 nM | 72 | 76.2 ± 16, p ≤ 0.001 | 102.7 ± 24.4, p = 0.62 | 57.6 ± 14.7, p ≤ 0.001 | 98.2 ± 20.5, p = 0.2 |
| High Risk | Salbutamol | 100 nM | 72 | 62.9 ± 15.7, p = 0.001 | 92.6 ± 24.3, p = 0.44 | 48.3 ± 13.3, p ≤ 0.001 | 97.6 ± 26, p = 0.33 |
| High Risk | Salbutamol | 300 nM | 72 | 56.5 ± 13.5, p ≤ 0.001 | 87.8 ± 20.9, p = 0.97 | 43.5 ± 12, p ≤ 0.001 | 94.4 ± 26.1, p = 0.21 |
| Low Risk | Tetrodotoxin | 100 nM | 49 | 107.1 ± 4.9, p = 0.02 | 106.4 ± 5, p = 0.48 | 101.6 ± 7.4, p = 0.27 | 103.1 ± 41.9, p = 0.08 |
| Low Risk | Tetrodotoxin | 300 nM | 49 | 110.3 ± 6.8, p = 0.05 | 108.7 ± 11.2, p = 0.44 | 108.6 ± 52.8, p = 0.002 | 93.6 ± 38, p = 0.2 |
| Low Risk | Tetrodotoxin | 1 µM | 49 | 111.4 ± 7.6, p = 0.15 | 109.3 ± 8, p = 0.77 | 104.6 ± 13.5, p = 0.05 | 74.4 ± 30.6, p = 0.88 |
| Low Risk | Tetrodotoxin | 10 µM | 49 | 109.7 ± 9.3, p = 0.84 | 101.2 ± 10.7, p = 0.59 | 121.2 ± 31.4, p = 0.3 | 45.9 ± 22.3, p = 0.98 |
| Low Risk | Tetrodotoxin | 30 µM | 49 | 102 ± 7.8, p = 0.04 | 84.8 ± 10.3, p = 0.55 | 168.9 ± 87.5, p = 0.71 | 9.5 ± 8.8, p = 0.003 |
| High Risk | Tetrodotoxin | 100 nM | 72 | 105 ± 4, p = 0.02 | 106.4 ± 7.8, p = 0.48 | 98.6 ± 13.9, p = 0.27 | 95.5 ± 18.3, p = 0.08 |
| High Risk | Tetrodotoxin | 300 nM | 72 | 107.5 ± 7, p = 0.05 | 111.1 ± 10.9, p = 0.44 | 95.3 ± 16.4, p = 0.002 | 86.4 ± 19.8, p = 0.2 |
| High Risk | Tetrodotoxin | 1 µM | 72 | 108.1 ± 8.5, p = 0.15 | 109.6 ± 13.2, p = 0.77 | 101.6 ± 28.8, p = 0.05 | 70.6 ± 20.3, p = 0.88 |
| High Risk | Tetrodotoxin | 10 µM | 72 | 108.7 ± 9.3, p = 0.84 | 103.6 ± 18.6, p = 0.59 | 127.9 ± 92.6, p = 0.3 | 44.5 ± 19, p = 0.98 |
| High Risk | Tetrodotoxin | 30 µM | 69 | 108.7 ± 12.2, p = 0.04 | 92 ± 27.1, p = 0.55 | 160.4 ± 60.5, p = 0.71 | 19.2 ± 13.5, p = 0.003 |

Changes of field potential duration (FPD) corrected field potential duration (cFPD), RR interval (RR), and peak-to-peak amplitude (PtPA) are presented as percentage of the Vehicle treatment response. Values are presented as mean ± standard deviation. N indicates the number of hiPSC-CMs monolayers included in the analysis.

**Supplemental Table 8. Differential drug-concentration responses to dofetilide, E4031, and haloperidol in High Risk and Low Risk *KCNQ1* hiPSC-CMs.**

| **Variant Risk Level** | **Drug** | **Dose Label** | **N** | **Normalized FPD (% of Vehicle)** | **Normalized cFPD (% of Vehicle)** | **Normalized RR (% of Vehicle)** | **Normalized PtPA (% of Vehicle)** |
| --- | --- | --- | --- | --- | --- | --- | --- |
| Low Risk | Dofetilide | 1 nM | 17 | 108.8 ± 2.6, p = 0.08 | 109.2 ± 2.8, p = 0.1 | 99.3 ± 4.2, p = 0.7 | 100.4 ± 16.8, p = 0.87 |
| Low Risk | Dofetilide | 3 nM | 17 | 125.9 ± 7.2, p = 0.09 | 126.2 ± 7.3, p = 0.02 | 99.7 ± 5.4, p = 0.43 | 102.1 ± 16.6, p = 0.06 |
| Low Risk | Dofetilide | 10 nM | 17 | 172.1 ± 23.8, p = 0.05 | 166.1 ± 21.7, p = 0.01 | 107.4 ± 7.4, p = 0.71 | 95.3 ± 22.4, p = 0.07 |
| Low Risk | Dofetilide | 30 nM | 17 | 233.3 ± 71.9, p = 0.005 | 206.2 ± 40, p ≤ 0.001 | 129.8 ± 37.6, p = 0.85 | 84.2 ± 23.5, p = 0.25 |
| Low Risk | Dofetilide | 100 nM | 17 | 251.5 ± 159.5, p ≤ 0.001 | 218.7 ± 88.9, p ≤ 0.001 | 182.7 ± 108.9, p = 0.33 | 59.9 ± 33.2, p = 0.66 |
| High Risk | Dofetilide | 1 nM | 101 | 111.3 ± 7, p = 0.08 | 112.3 ± 10.3, p = 0.1 | 100.1 ± 17, p = 0.7 | 99 ± 8.5, p = 0.87 |
| High Risk | Dofetilide | 3 nM | 101 | 131.5 ± 15.4, p = 0.09 | 134.8 ± 16.7, p = 0.02 | 96.9 ± 17.9, p = 0.43 | 95.5 ± 13.9, p = 0.06 |
| High Risk | Dofetilide | 10 nM | 101 | 197.5 ± 51.4, p = 0.05 | 191 ± 44.6, p = 0.01 | 127.5 ± 211, p = 0.71 | 87.2 ± 18.6, p = 0.07 |
| High Risk | Dofetilide | 30 nM | 101 | 303.8 ± 110.5, p = 0.005 | 274.4 ± 100, p ≤ 0.001 | 137.4 ± 84.4, p = 0.85 | 74.6 ± 25.8, p = 0.25 |
| High Risk | Dofetilide | 100 nM | 101 | 377.8 ± 154.6, p ≤ 0.001 | 319.2 ± 129.6, p ≤ 0.001 | 165.4 ± 150.2, p = 0.33 | 58 ± 31, p = 0.66 |
| Low Risk | E4031 | 10 nM | 16 | 121.1 ± 18, p = 0.69 | 119.6 ± 15.1, p = 0.44 | 102.4 ± 8, p = 0.62 | 98.7 ± 16.3, p = 0.3 |
| Low Risk | E4031 | 30 nM | 16 | 139.9 ± 22.9, p ≤ 0.001 | 139 ± 18, p ≤ 0.001 | 100.7 ± 8.6, p = 0.19 | 101.8 ± 18.2, p = 0.002 |
| Low Risk | E4031 | 100 nM | 16 | 172.4 ± 32.9, p ≤ 0.001 | 168.7 ± 23.9, p ≤ 0.001 | 103.5 ± 11.3, p = 0.13 | 92.9 ± 16.1, p = 0.004 |
| Low Risk | E4031 | 300 nM | 16 | 193.6 ± 30.9, p ≤ 0.001 | 185.9 ± 19.1, p ≤ 0.001 | 108.9 ± 23.9, p = 0.002 | 74.7 ± 33.3, p = 0.07 |
| Low Risk | E4031 | 500 nM | 16 | 199.4 ± 24.6, p ≤ 0.001 | 184.1 ± 37.4, p ≤ 0.001 | 167.4 ± 241, p = 0.08 | 66.2 ± 34.1, p = 0.15 |
| High Risk | E4031 | 10 nM | 64 | 129.6 ± 35.8, p = 0.69 | 128.6 ± 29, p = 0.44 | 101.1 ± 14, p = 0.62 | 97.6 ± 13.3, p = 0.3 |
| High Risk | E4031 | 30 nM | 64 | 199.4 ± 80.1, p ≤ 0.001 | 192.6 ± 82.8, p ≤ 0.001 | 109.8 ± 25.5, p = 0.19 | 88.7 ± 15.1, p = 0.002 |
| High Risk | E4031 | 100 nM | 64 | 291.1 ± 200.1, p ≤ 0.001 | 256.5 ± 138.6, p ≤ 0.001 | 122.8 ± 46.5, p = 0.13 | 72.6 ± 26.4, p = 0.004 |
| High Risk | E4031 | 300 nM | 64 | 394.6 ± 225.9, p ≤ 0.001 | 333 ± 173, p ≤ 0.001 | 147 ± 74, p = 0.002 | 61.9 ± 22.7, p = 0.07 |
| High Risk | E4031 | 500 nM | 60 | 447.1 ± 250.4, p ≤ 0.001 | 377.2 ± 184.9, p ≤ 0.001 | 171 ± 180.2, p = 0.08 | 52.9 ± 27.4, p = 0.15 |
| Low Risk | Haloperidol | 300 nM | 16 | 134.6 ± 30.9, p = 0.009 | 124 ± 24.7, p = 0.002 | 117.7 ± 19.9, p = 0.56 | 96.5 ± 25.1, p = 0.84 |
| Low Risk | Haloperidol | 1 µM | 16 | 220.2 ± 87.5, p = 0.006 | 183.4 ± 50.7, p = 0.002 | 142.1 ± 52.2, p = 0.33 | 71.1 ± 24.6, p = 0.39 |
| Low Risk | Haloperidol | 3 µM | 16 | 221.6 ± 75.8, p = 0.008 | 180.4 ± 39.1, p = 0.001 | 151.1 ± 61.8, p = 0.69 | 42.6 ± 30.4, p = 0.2 |
| Low Risk | Haloperidol | 10 µM | 16 | 192.5 ± 54.1, p = 0.39 | 154.4 ± 25.6, p = 0.007 | 169.6 ± 87, p = 0.04 | 31.7 ± 26.2, p = 0.7 |
| High Risk | Haloperidol | 300 nM | 99 | 203.3 ± 151.2, p = 0.009 | 182.5 ± 116.6, p = 0.002 | 120.3 ± 31.4, p = 0.56 | 92.8 ± 14.5, p = 0.84 |
| High Risk | Haloperidol | 1 µM | 99 | 312 ± 192.3, p = 0.006 | 258.7 ± 142.7, p = 0.002 | 149 ± 68.3, p = 0.33 | 75.3 ± 23.9, p = 0.39 |
| High Risk | Haloperidol | 3 µM | 99 | 301.9 ± 154.3, p = 0.008 | 264 ± 129.6, p = 0.001 | 143.8 ± 62.2, p = 0.69 | 50.5 ± 27.1, p = 0.2 |
| High Risk | Haloperidol | 10 µM | 99 | 252.6 ± 175.7, p = 0.39 | 226 ± 129.7, p = 0.007 | 153.4 ± 138.4, p = 0.04 | 34.5 ± 26.5, p = 0.7 |

Changes of field potential duration (FPD) corrected field potential duration (cFPD), RR interval (RR), and peak-to-peak amplitude (PtPA) are presented as percentage of the Vehicle treatment response. Values are presented as mean ± standard deviation. N indicates the number of hiPSC-CMs monolayers included in the analysis.

**Supplemental Table 9. Differential drug-concentration responses to dofetilide, E4031, and haloperidol in High Risk and Low Risk *KCNH2* hiPSC-CMs.**

| **Variant Risk Level** | **Drug** | **Dose Label** | **N** | **Normalized FPD (% of Vehicle)** | **Normalized cFPD (% of Vehicle)** | **Normalized RR (% of Vehicle)** | **Normalized PtPA (% of Vehicle)** |
| --- | --- | --- | --- | --- | --- | --- | --- |
| Low Risk | Dofetilide | 1 nM | 30 | 110.1 ± 3.8, p = 0.59 | 108.1 ± 5, p = 0.4 | 104.3 ± 8.2, p = 0.009 | 97.8 ± 10.7, p = 0.09 |
| Low Risk | Dofetilide | 3 nM | 30 | 125.3 ± 8.2, p = 0.18 | 123.1 ± 7.8, p = 0.004 | 104.2 ± 11, p = 0.59 | 94.9 ± 17.8, p = 0.25 |
| Low Risk | Dofetilide | 10 nM | 30 | 157.5 ± 20.8, p ≤ 0.001 | 150.5 ± 17.9, p ≤ 0.001 | 111.3 ± 23.3, p = 0.27 | 84.5 ± 27.9, p = 0.08 |
| Low Risk | Dofetilide | 30 nM | 30 | 202.4 ± 46.1, p = 0.71 | 189.6 ± 41.6, p = 0.71 | 114.9 ± 18.3, p = 0.09 | 77.1 ± 26.6, p ≤ 0.001 |
| Low Risk | Dofetilide | 100 nM | 30 | 236.3 ± 77.2, p = 0.09 | 216.6 ± 64.7, p = 0.1 | 126.2 ± 41.8, p = 0.07 | 67.6 ± 23.6, p = 0.002 |
| High Risk | Dofetilide | 1 nM | 20 | 110.4 ± 6.7, p = 0.59 | 110.2 ± 7.3, p = 0.4 | 101.1 ± 12.1, p = 0.009 | 103.2 ± 9.6, p = 0.09 |
| High Risk | Dofetilide | 3 nM | 20 | 141.7 ± 31.9, p = 0.18 | 138.3 ± 21.4, p = 0.004 | 104.3 ± 19.3, p = 0.59 | 95.7 ± 22.8, p = 0.25 |
| High Risk | Dofetilide | 10 nM | 20 | 231.9 ± 71.8, p ≤ 0.001 | 198.9 ± 50.1, p ≤ 0.001 | 131.7 ± 38.3, p = 0.27 | 69.4 ± 28.5, p = 0.08 |
| High Risk | Dofetilide | 30 nM | 20 | 255.1 ± 146.9, p = 0.71 | 226.1 ± 111.5, p = 0.71 | 107 ± 51.3, p = 0.09 | 37.2 ± 34.1, p ≤ 0.001 |
| High Risk | Dofetilide | 100 nM | 20 | 331.7 ± 146, p = 0.09 | 297.4 ± 121.2, p = 0.1 | 100.2 ± 47.6, p = 0.07 | 33.5 ± 33.6, p = 0.002 |
| Low Risk | E4031 | 10 nM | 18 | 123.1 ± 19.5, p = 0.39 | 124.4 ± 16.5, p = 0.31 | 98.3 ± 13, p = 0.17 | 109.9 ± 28.7, p = 0.02 |
| Low Risk | E4031 | 30 nM | 18 | 172.7 ± 25.7, p = 0.13 | 171.2 ± 26.1, p = 0.39 | 103.1 ± 17.2, p = 0.004 | 98.9 ± 45.6, p = 0.42 |
| Low Risk | E4031 | 100 nM | 18 | 228.4 ± 54.9, p = 0.03 | 222.2 ± 58.6, p = 0.08 | 107.5 ± 17.1, p = 0.15 | 93.4 ± 60.6, p = 0.17 |
| Low Risk | E4031 | 300 nM | 18 | 274.1 ± 75.2, p = 0.45 | 259.9 ± 72, p = 0.37 | 112.7 ± 18.5, p = 0.05 | 86.5 ± 68.6, p = 0.05 |
| Low Risk | E4031 | 500 nM | 18 | 283.1 ± 83.6, p = 0.78 | 269 ± 83.5, p = 0.13 | 113.1 ± 21.3, p = 0.7 | 82.9 ± 73.1, p = 0.002 |
| High Risk | E4031 | 10 nM | 13 | 122 ± 31, p = 0.39 | 118 ± 24.2, p = 0.31 | 105.5 ± 12.6, p = 0.17 | 94.3 ± 8.1, p = 0.02 |
| High Risk | E4031 | 30 nM | 13 | 226.5 ± 80.8, p = 0.13 | 192.2 ± 56.2, p = 0.39 | 135.9 ± 31, p = 0.004 | 81.6 ± 20.9, p = 0.42 |
| High Risk | E4031 | 100 nM | 13 | 286.7 ± 70.5, p = 0.03 | 238.6 ± 49.5, p = 0.08 | 132.2 ± 47.8, p = 0.15 | 63.5 ± 31.9, p = 0.17 |
| High Risk | E4031 | 300 nM | 13 | 275.3 ± 118.8, p = 0.45 | 221.9 ± 86.1, p = 0.37 | 153.1 ± 70.5, p = 0.05 | 46.5 ± 32.6, p = 0.05 |
| High Risk | E4031 | 500 nM | 13 | 242.8 ± 114.3, p = 0.78 | 211 ± 90.7, p = 0.13 | 123.8 ± 64.3, p = 0.7 | 27.3 ± 32.4, p = 0.002 |
| Low Risk | Haloperidol | 300 nM | 27 | 139 ± 38.3, p = 0.02 | 129.6 ± 31.9, p = 0.009 | 115.2 ± 22.6, p = 0.01 | 89.9 ± 24.1, p = 0.34 |
| Low Risk | Haloperidol | 1 µM | 27 | 220.8 ± 78.3, p = 0.005 | 189.3 ± 57.4, p = 0.04 | 139.3 ± 43.4, p = 0.41 | 64.6 ± 37.5, p = 0.29 |
| Low Risk | Haloperidol | 3 µM | 27 | 212.9 ± 56.9, p = 0.45 | 192.8 ± 39.2, p = 0.11 | 122.4 ± 29.5, p = 0.82 | 50.5 ± 38.5, p = 0.1 |
| Low Risk | Haloperidol | 10 µM | 27 | 163.8 ± 38, p = 0.02 | 161.7 ± 39.4, p = 0.009 | 104.1 ± 21.5, p = 1 | 52.8 ± 37.9, p = 0.003 |
| High Risk | Haloperidol | 300 nM | 19 | 203.8 ± 92.8, p = 0.02 | 167.8 ± 55.3, p = 0.009 | 140 ± 38.6, p = 0.01 | 86 ± 22.4, p = 0.34 |
| High Risk | Haloperidol | 1 µM | 19 | 275.8 ± 64.8, p = 0.005 | 218.2 ± 43.7, p = 0.04 | 150.2 ± 51, p = 0.41 | 54.4 ± 30.7, p = 0.29 |
| High Risk | Haloperidol | 3 µM | 19 | 194.7 ± 38.3, p = 0.45 | 169.8 ± 32.8, p = 0.11 | 118.9 ± 40.9, p = 0.82 | 31.7 ± 28.5, p = 0.1 |
| High Risk | Haloperidol | 10 µM | 19 | 132.9 ± 33.5, p = 0.02 | 125.4 ± 29.3, p = 0.009 | 99.5 ± 31, p = 1 | 21.7 ± 23.3, p = 0.003 |

Changes of field potential duration (FPD) corrected field potential duration (cFPD), RR interval (RR), and peak-to-peak amplitude (PtPA) are presented as percentage of the Vehicle treatment response. Values are presented as mean ± standard deviation. N indicates the number of hiPSC-CMs monolayers included in the analysis.

### **Supplemental figures**

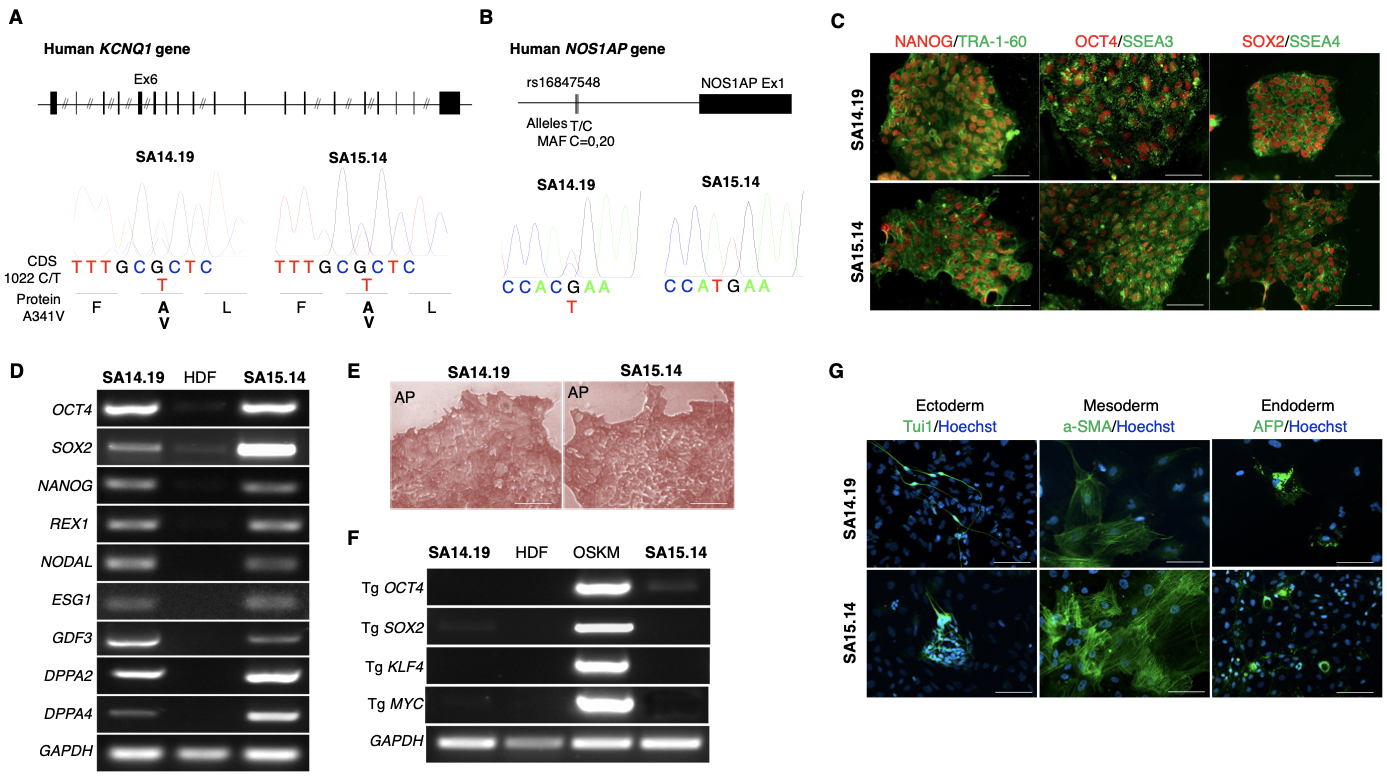

**Supplemental Figure 1. Characterization of SA14.19 and SA15.14 hiPSC lines.**

A. Top: schematic representation of KCNQ1 gene (exons are vertical lines/boxes). Bottom: DNA sequencing results showing the mutation 1022 C/T in the KCNQ1 exon 6 (Ex6) in heterozygosis in the both SA14.19 and SA15.14 hiPSCs. B. Top: schematic representation of *NOS1AP* gene upstream region. MAF is the minor allele frequency in the SA founder population. Bottom: DNA sequencing results showing the rs16847548 minor allele in heterozygosis in SA14.19 but not in SA15.14 hiPSCs. C. Immunofluorescence stainings showing uniform expression of the indicated markers of pluripotency in the SA14.19 and SA15.14 hiPSCs. Nuclear transcription factors are in red, membrane antigens are in green. Scale bars 100 um. D. RT-PCR analysis showing induction of expression of the indicated markers of pluripotency in SA14.19 and SA15.14 hiPSCs compared with fibroblasts before reprogramming. E. Alkaline phosphatase colorimetric staining (AP). Scale bars 100 um. F. RT-PCR analysis showing no expression of the four viral transgenes (Tg) in naïve fibroblasts (HDF), expression of Tg OCT4, SOX2, KLF4 and cMYC five days after transduction (OSKM) and silencing of the four Tg in SA14.19 and SA15.14 at passages 12. G. Immunofluorescence staining for markers of the 3 germ layers in iPSC-derived EBs: neuronal class tubulin beta III (Tuj) for ectoderm, smooth muscle actin (SMA) for mesoderm, and alpha Fetoprotein (AFP) for endoderm. Scale bars 100 um.

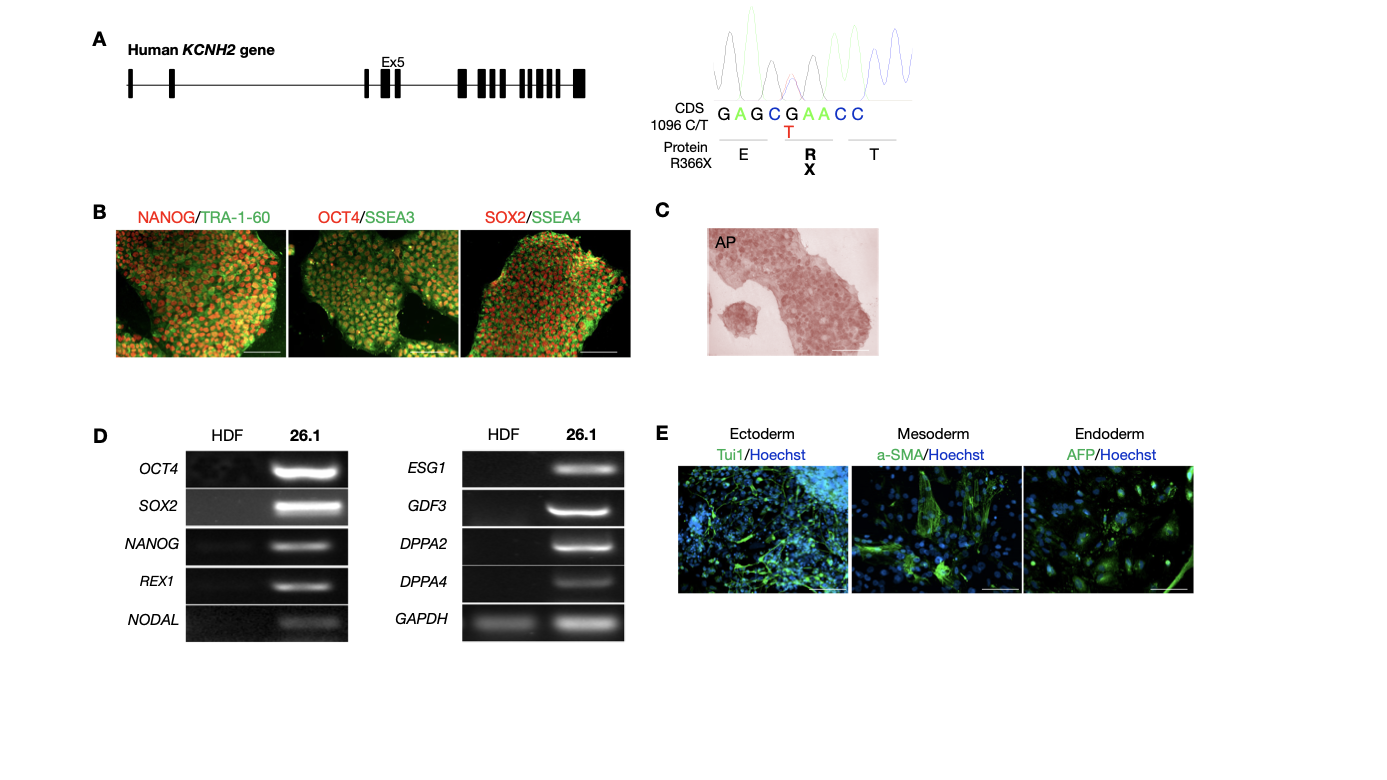

**Supplemental Figure 2. Characterization of 26.1 hiPSC line.**

A. Left: schematic representation of KCNH2 gene (exons are vertical boxes). Right: DNA sequencing results showing the mutation 1096 C/T in the KCNH2 exon 5 (Ex5) in heterozygosis in 26.1 hiPSCs. B. Immunofluorescence stainings showing uniform expression of the indicated markers of pluripotency in the 26.1 hiPSCs. Nuclear transcription factors are in red, membrane antigens are in green. Scale bars 100 um. C. Alkaline phosphatase colorimetric staining (AP). Scale bars 100 um. D.RT-PCR analysis showing induction of expression of the indicated markers of pluripotency in 26.1 hiPSCs compared with parental fibroblasts. E. Immunofluorescence staining for markers of the three germ layers in iPSC-derived EBs: neuronal class tubulin beta III (Tuj) for ectoderm, smooth muscle actin (SMA) for mesoderm, and alpha Fetoprotein (AFP) for endoderm. Scale bars 100 um.

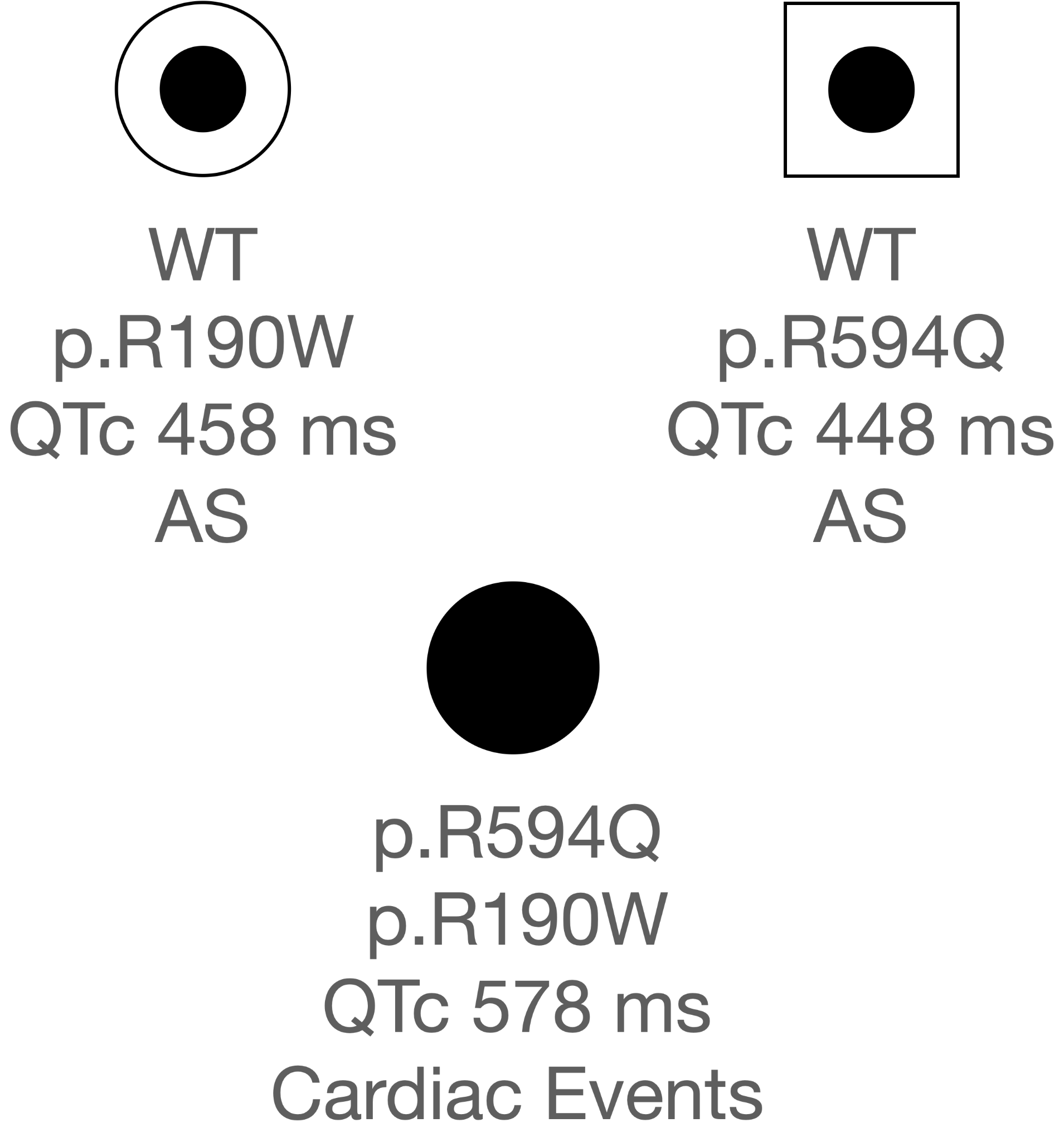

**Supplemental Figure 3.** Trio carrying genetic variants *KCNQ1* p.R190W and *KCNQ1* p.R594Q.

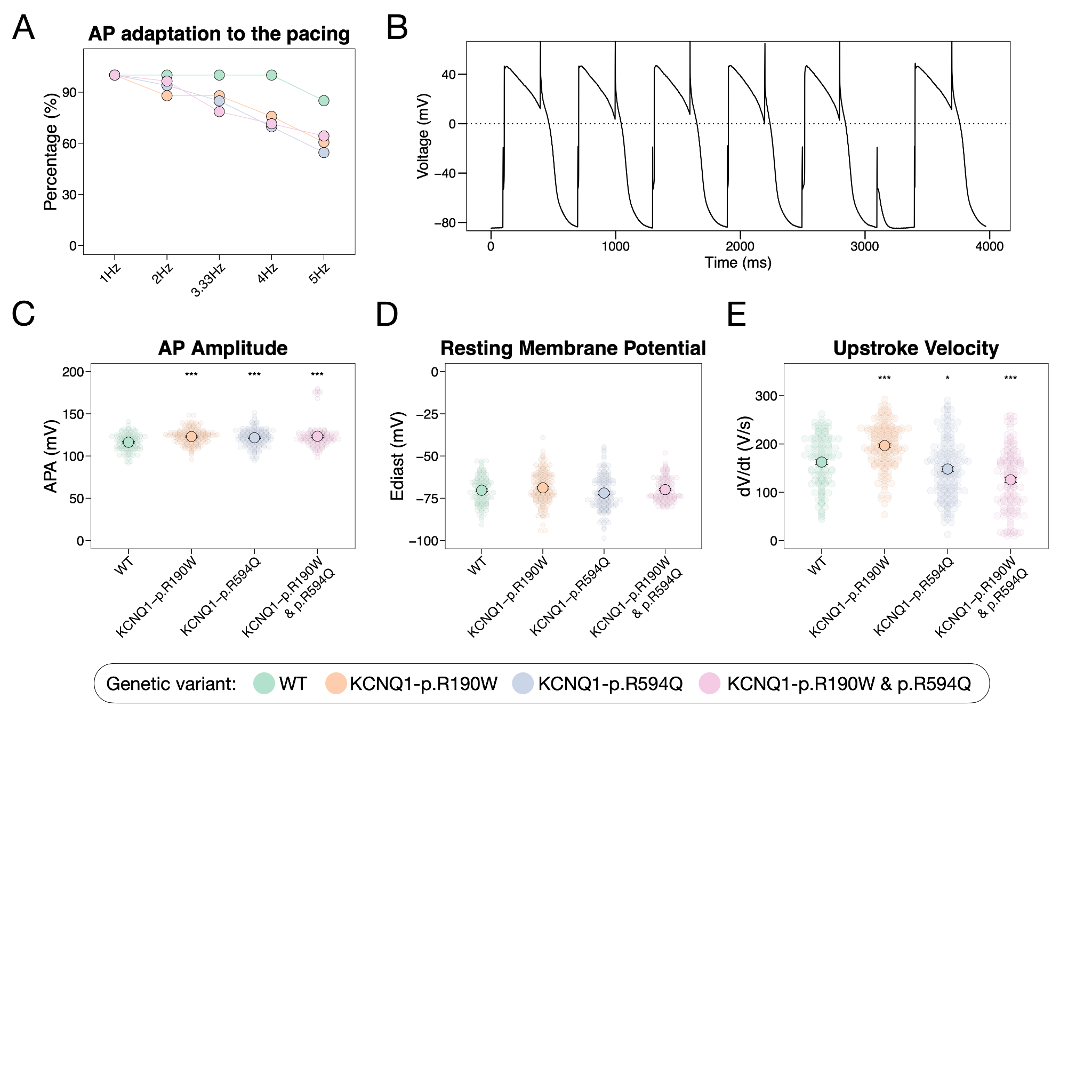

**Supplemental Figure 4. Characterization of hiPSC-CMs derived from family trio carrying genetic variants *KCNQ1* p.R190W and *KCNQ1* p.R594Q**

Color defines hiPSC-CMs carrying different genetic variants - green: wild type (WT), orange: *KCNQ1* p.R190W, blue: *KCNQ1* p.R594Q, and pink: *KCNQ1* p.R190W & p.R594Q. A. Percentage of hiPSC-CMs adapted to the increasing frequency pacing rate. B. Example of AP recording from hiPSC-CM carrying *KCNQ1* p.R190W & p.R594Q genetic variants not able to follow the 3 Hz pacing. C. hiPSC-CM action potential amplitude (APA). D. hiPSC-CM resting membrane potential. F. hiPSC-CM upstroke velocity.

**Supplemental Figure 5.** **Clarithromycin MEA dose-response curves of hiPSC-CMs from Low Risk and High Risk groups.**
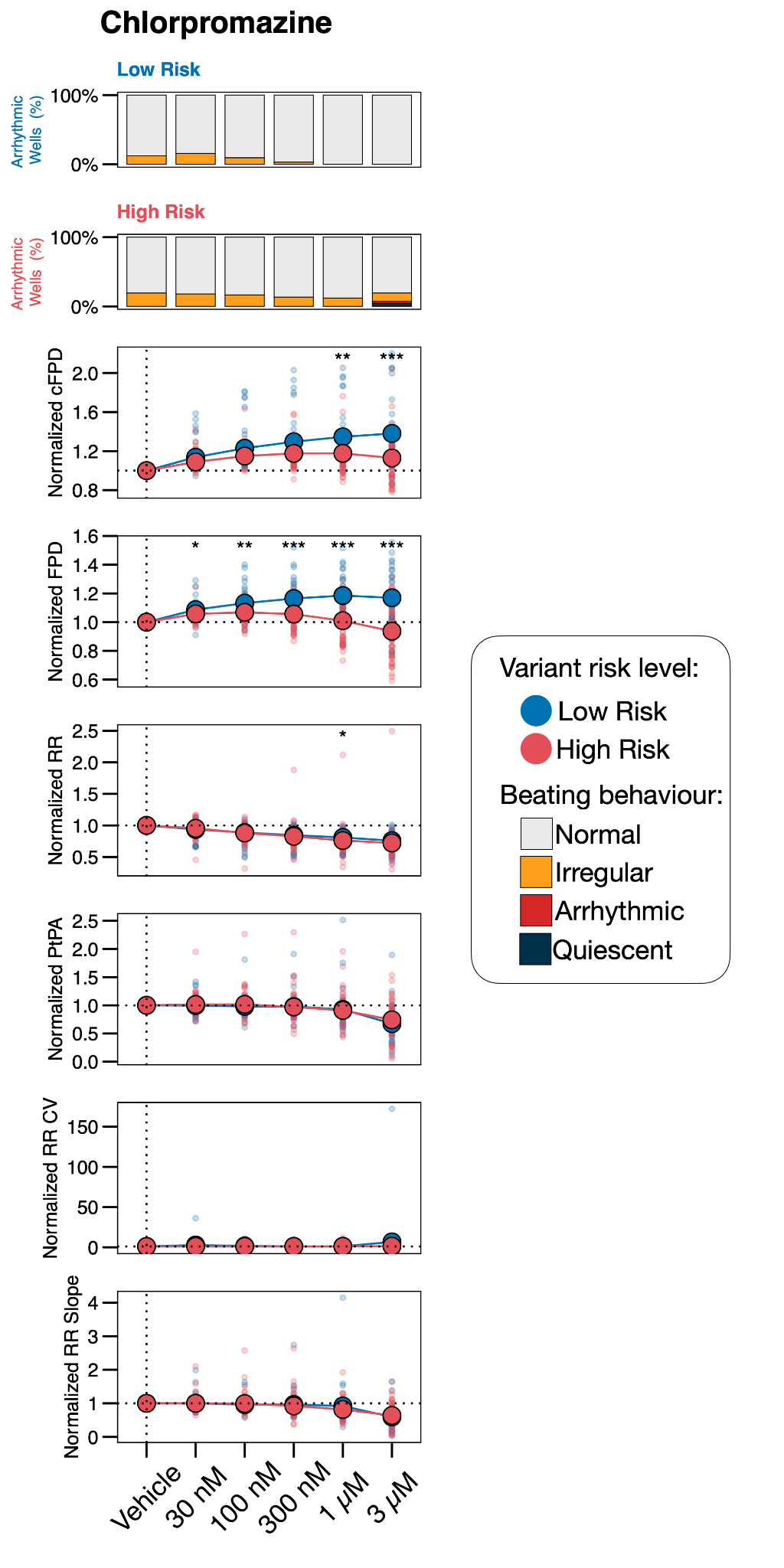

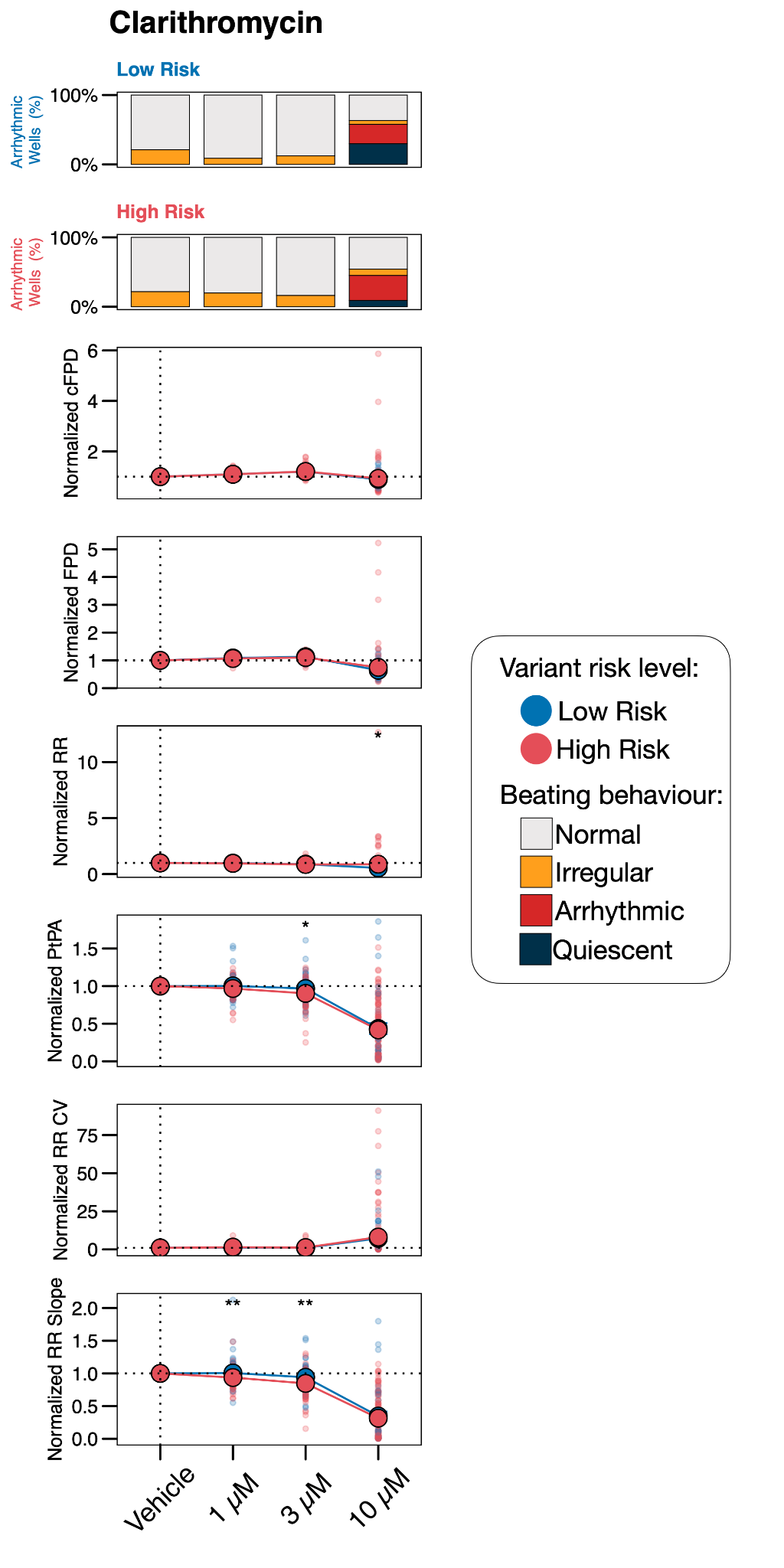

**Supplemental Figure 6.** **Chlorpromazine MEA dose-response curves of hiPSC-CMs from Low Risk and High Risk groups.**

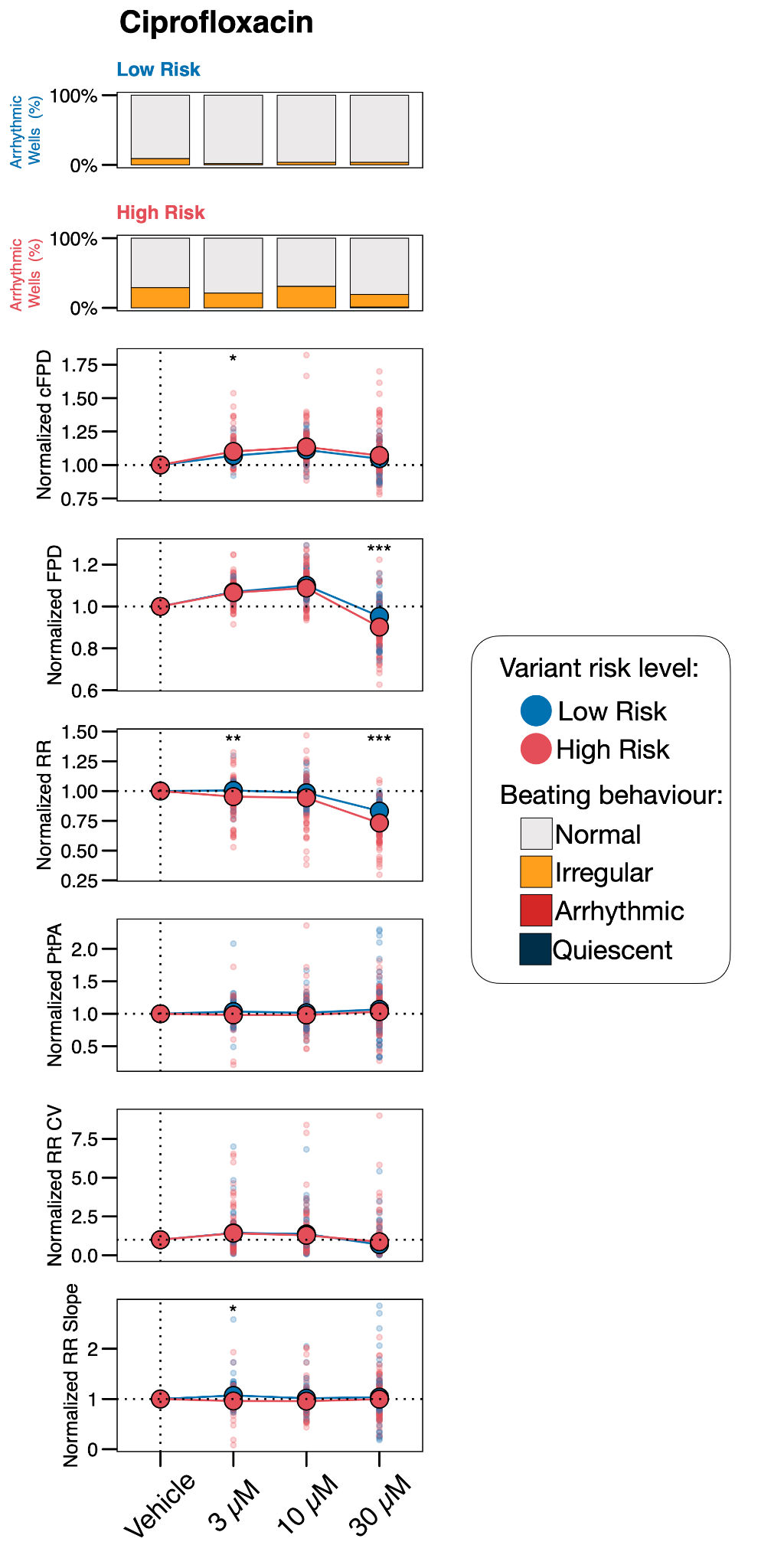

**Supplemental Figure 7.** **Ciprofloxacin MEA dose-response curves of hiPSC-CMs from Low Risk and High Risk groups.**

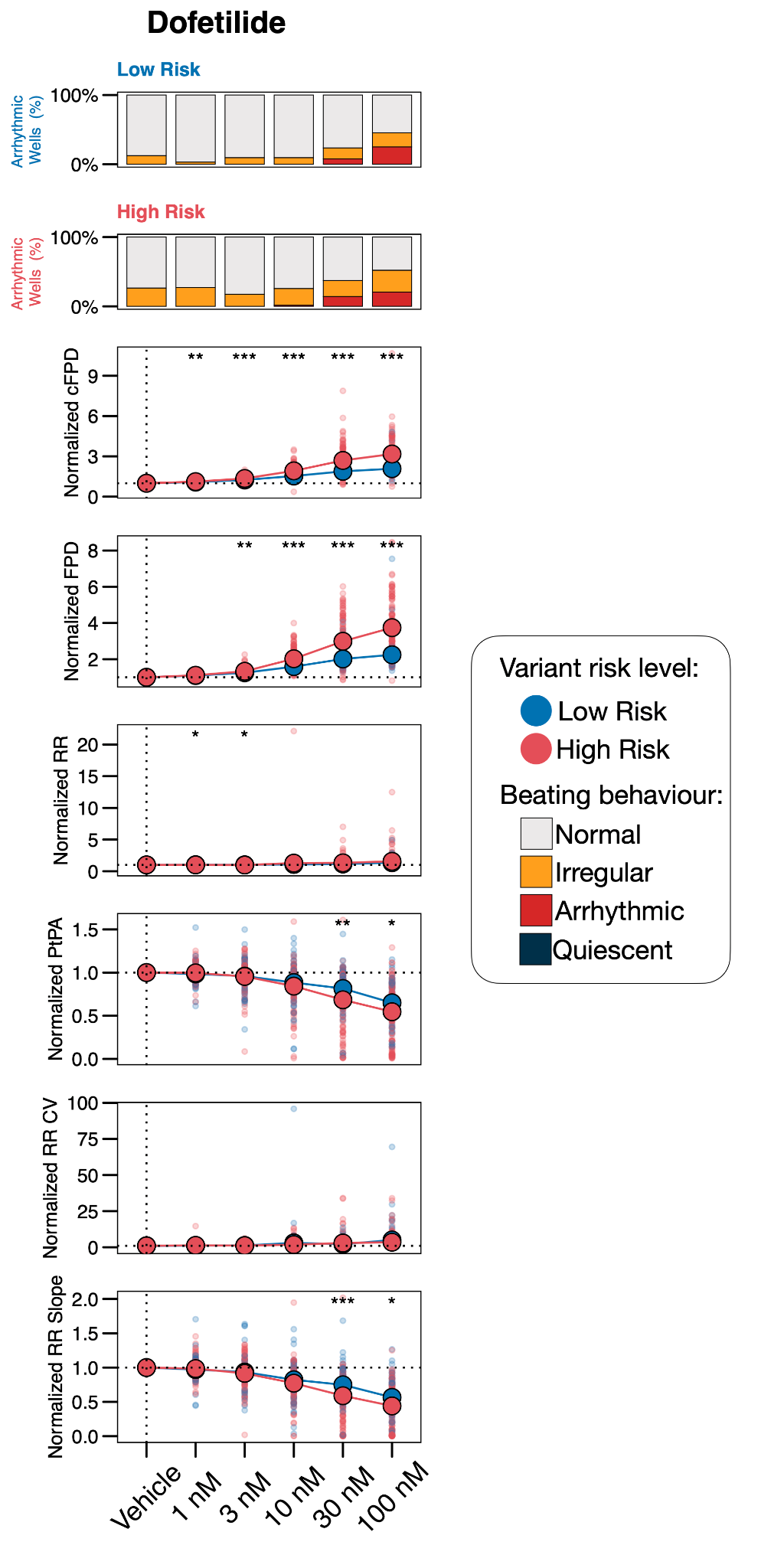

**Supplemental Figure 8.** **Dofetilide MEA dose-response curves of hiPSC-CMs from Low Risk and High Risk groups.**

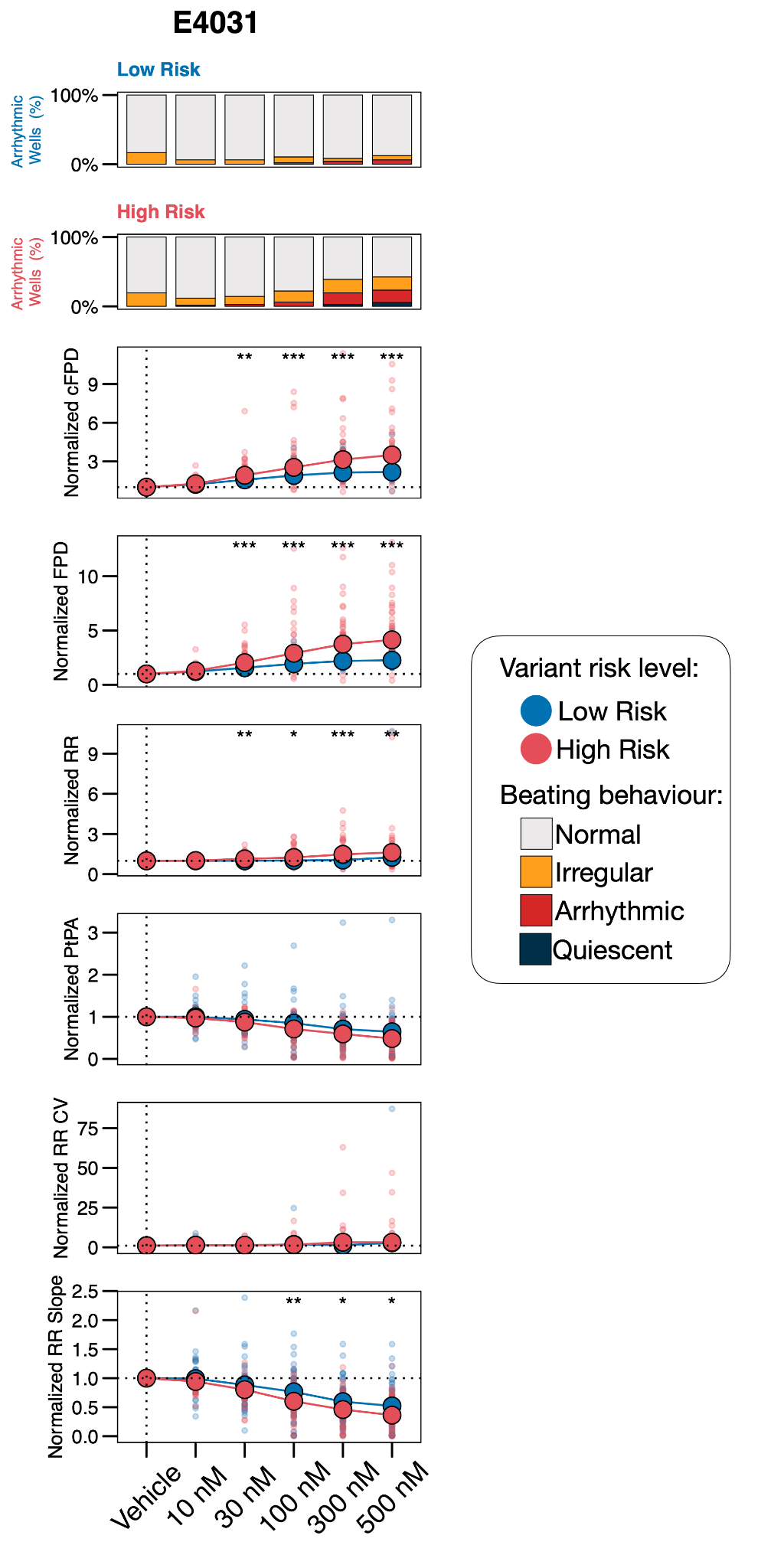

**Supplemental Figure 9.** **E4031 MEA dose-response curves of hiPSC-CMs from Low Risk and High Risk groups.**

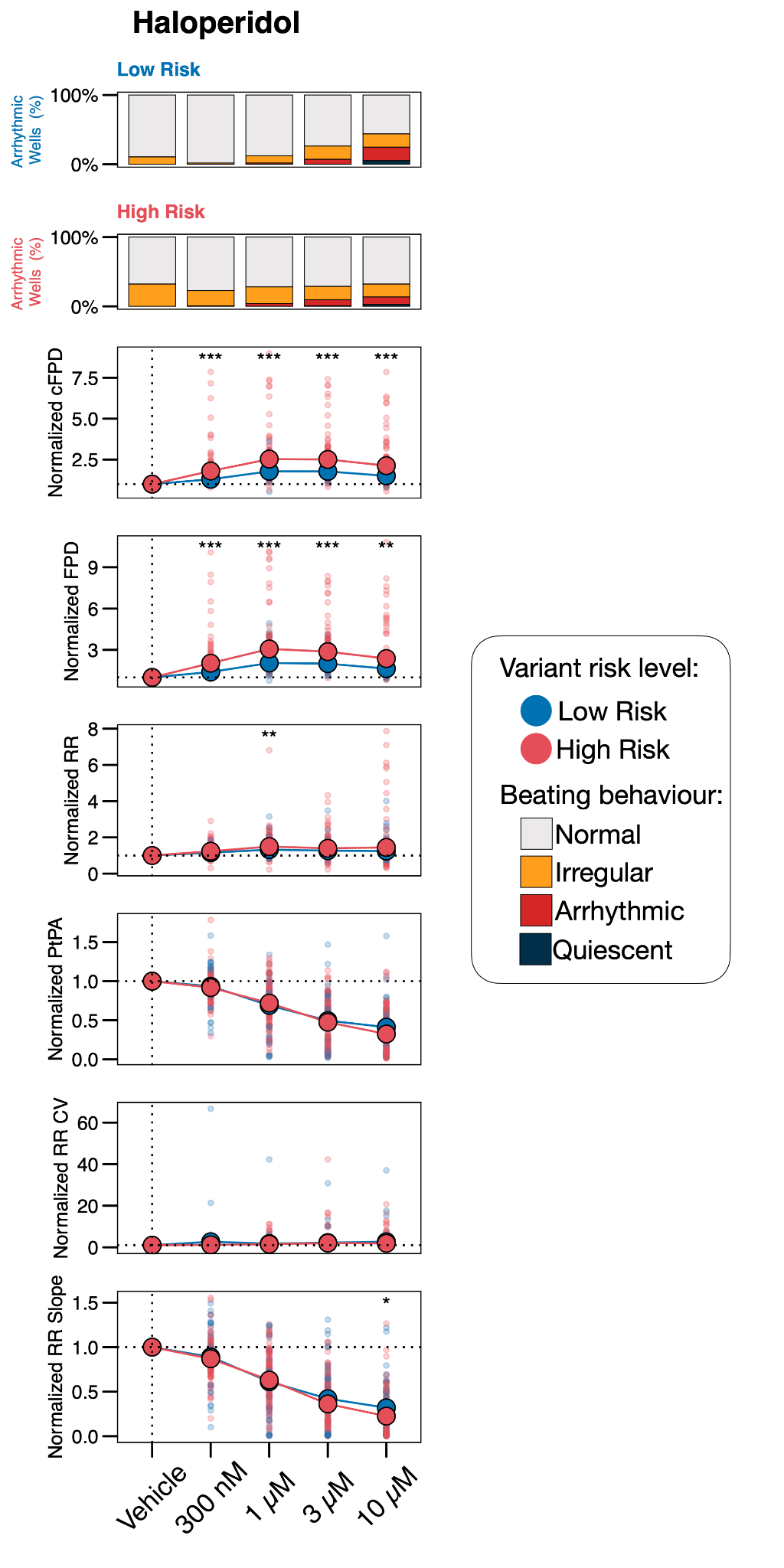

**Supplemental Figure 10.** **Haloperidol MEA dose-response curves of hiPSC-CMs from Low Risk and High Risk groups.**

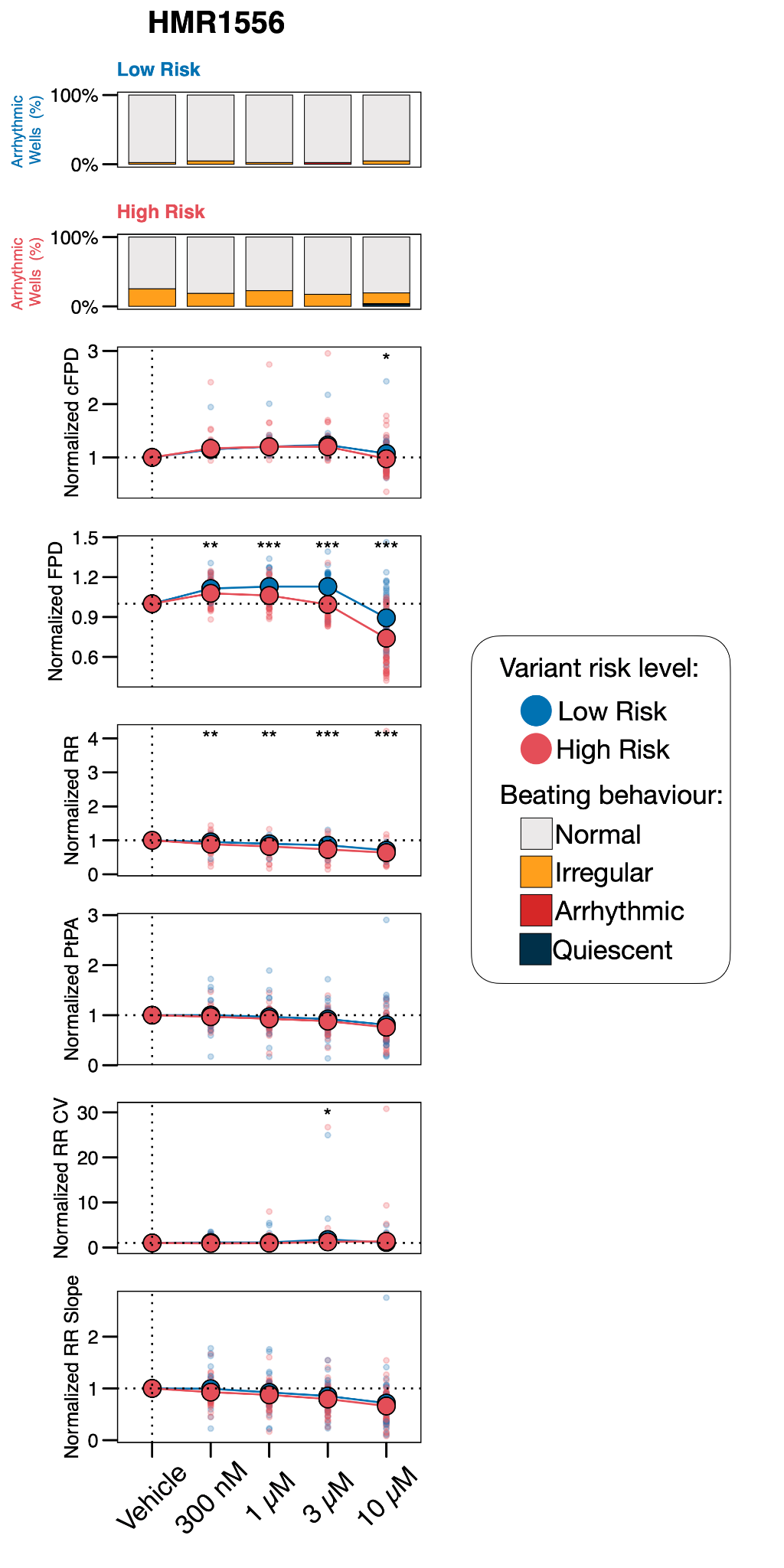

**Supplemental Figure 11.** **HMR1556 MEA dose-response curves of hiPSC-CMs from Low Risk and High Risk groups.**

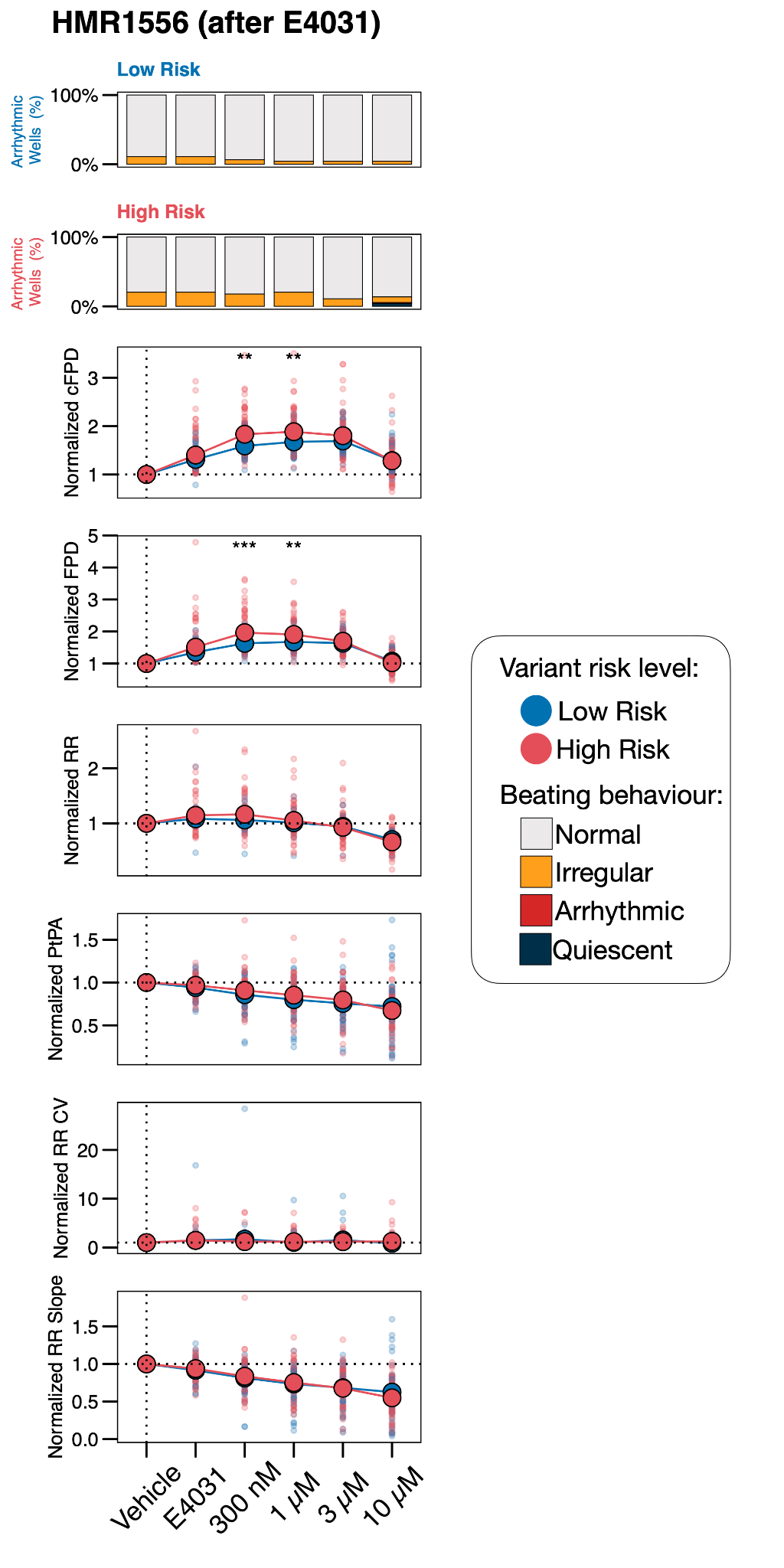

**Supplemental Figure 12.** **HMR1556 after E4031 pretreatment MEA dose-response curves of hiPSC-CMs from Low Risk and High Risk groups.**

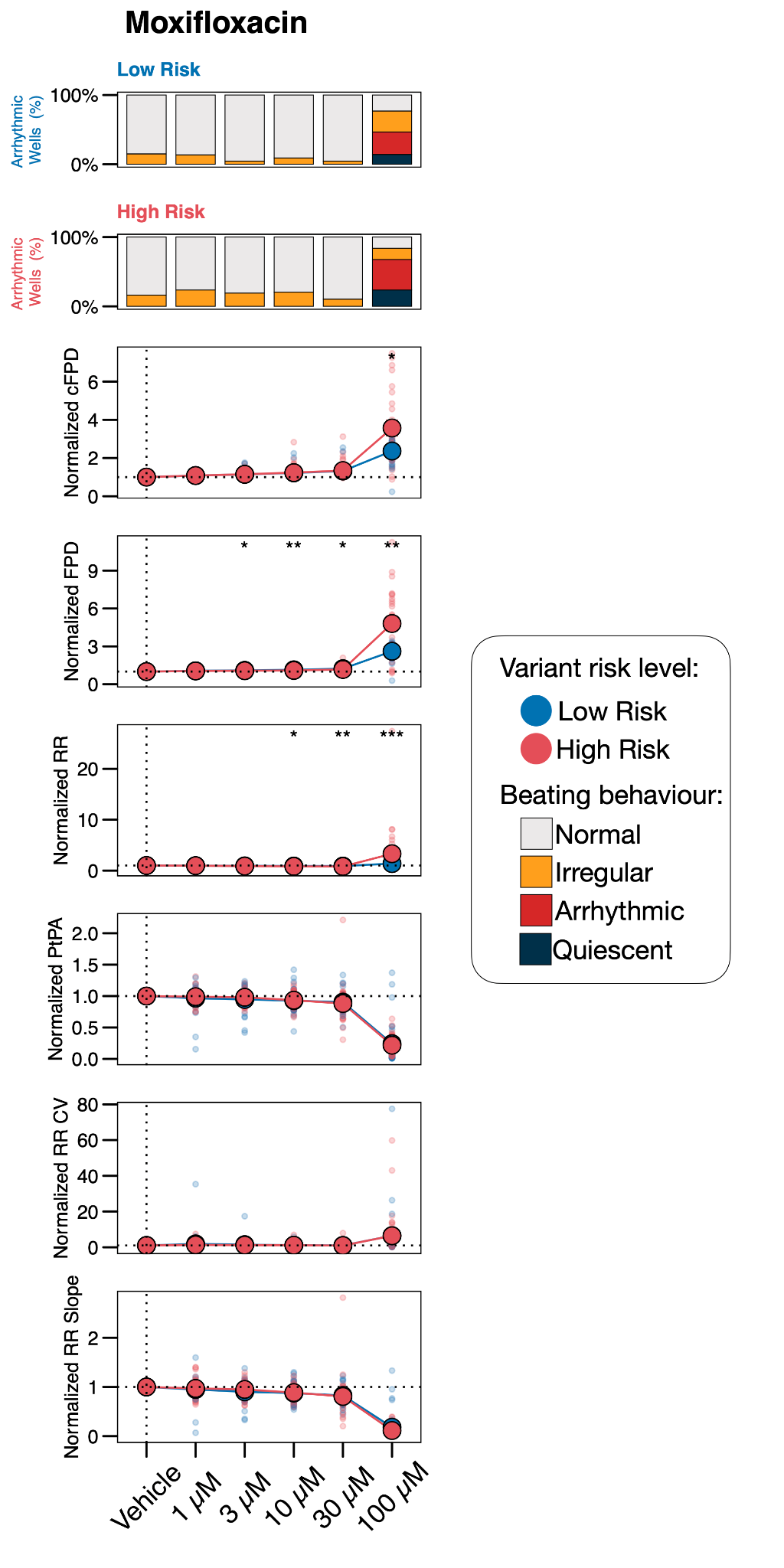

**Supplemental Figure 13.** **Moxifloxacin MEA dose-response curves of hiPSC-CMs from Low Risk and High Risk groups.**

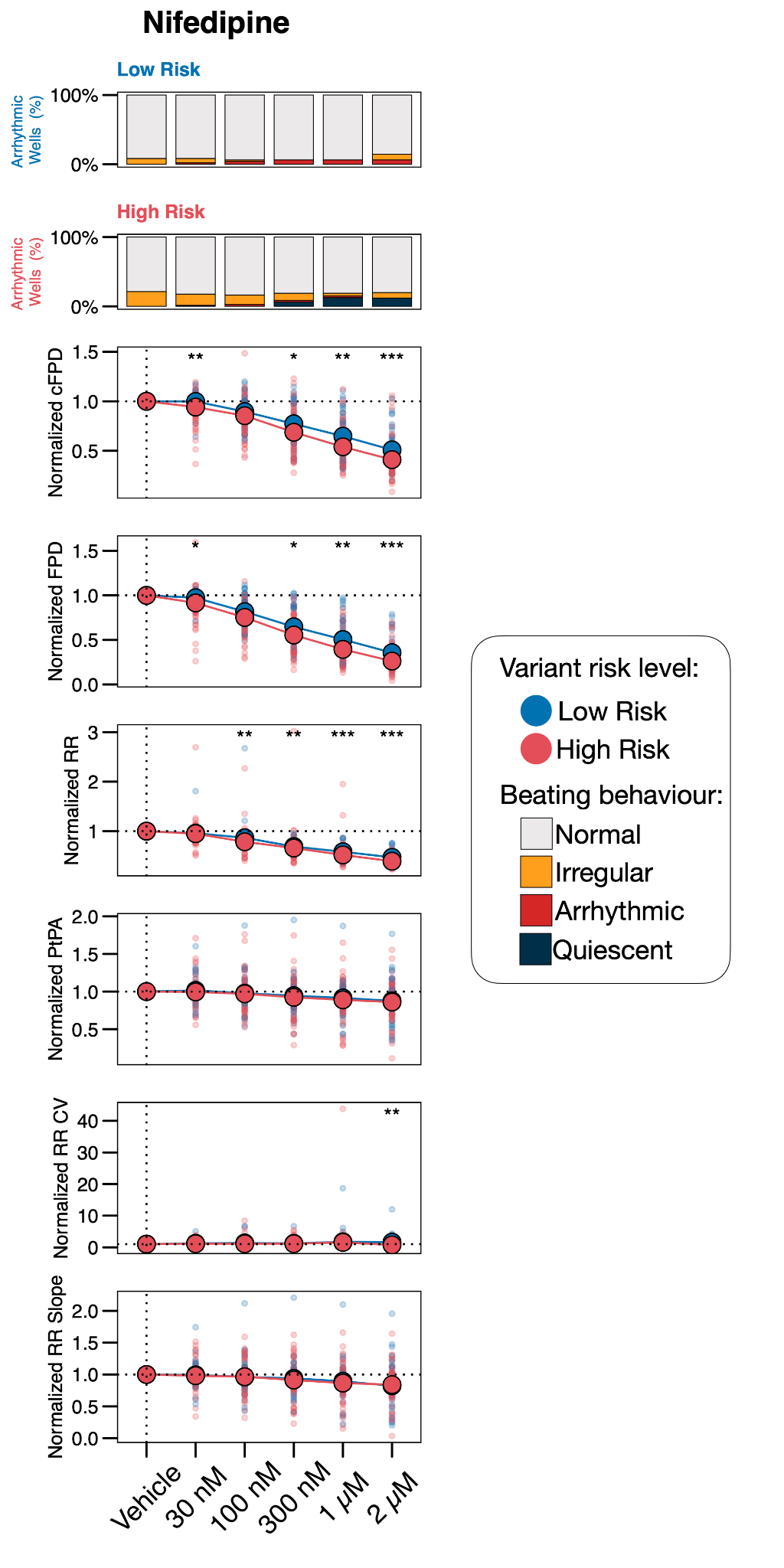

**Supplemental Figure 14.** **Nifedipine MEA dose-response curves of hiPSC-CMs from Low Risk and High Risk groups.**

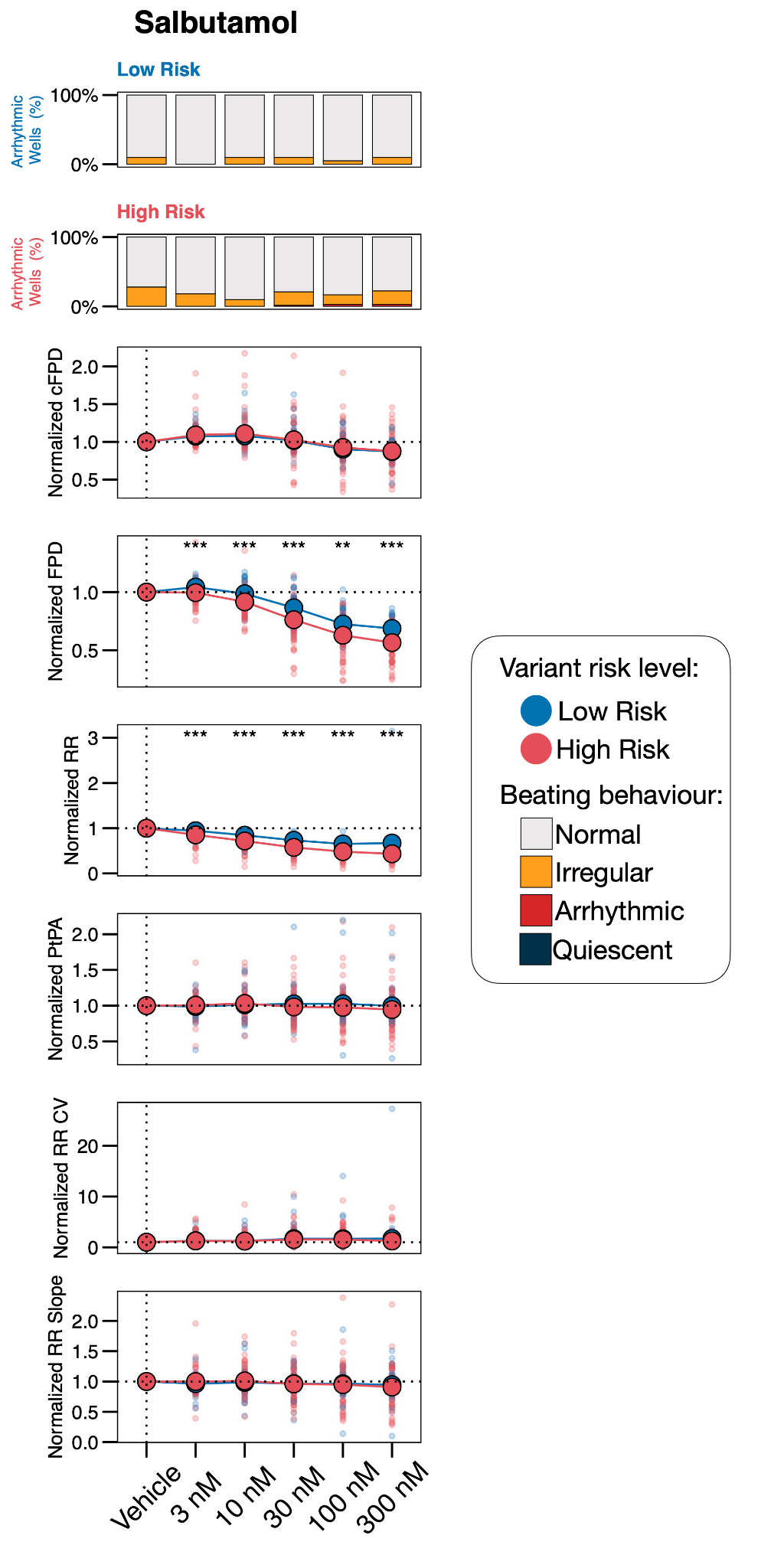

**Supplemental Figure 15.** **Salbutamol MEA dose-response curves of hiPSC-CMs from Low Risk and High Risk groups.**

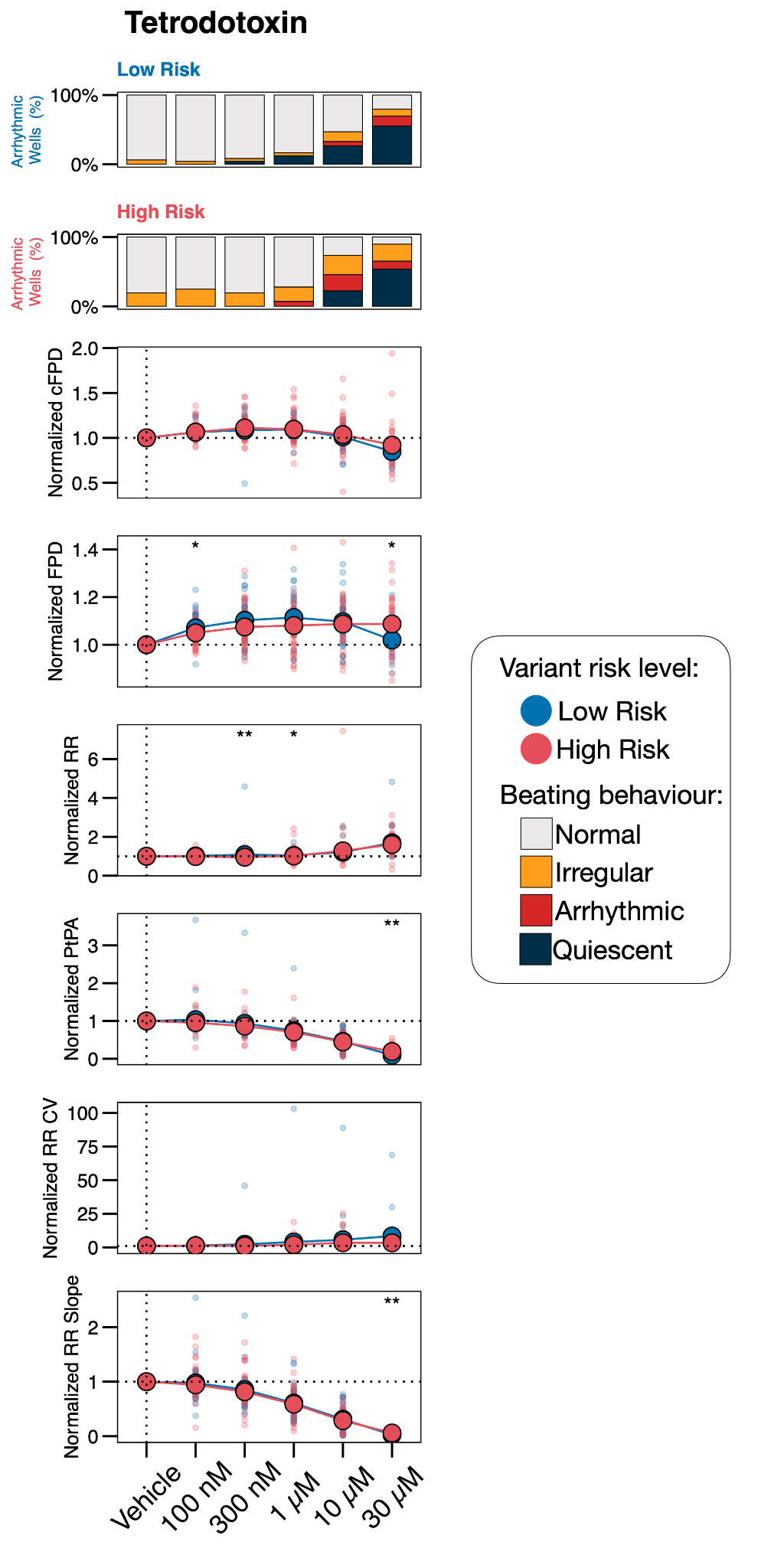
**3**

**Supplemental Figure 16.** **Tetrodotoxin MEA dose-response curves of hiPSC-CMs from Low Risk and High Risk groups.**
